# Automatic Extraction of Social Determinants of Health from Medical Notes of Chronic Lower Back Pain Patients

**DOI:** 10.1101/2022.03.04.22271541

**Authors:** Dmytro Lituiev, Benjamin Lacar, Sang Pak, Peter L Abramowitsch, Emilia De Marchis, Thomas Peterson

**Author notes:** Co-first authored (Dmytro Lituiev, Benjamin Lacar). Co-senior authored (Emilia De Marchis, Thomas Peterson).

## Abstract

**Background:** Adverse social determinants of health (SDoH), or social risk factors, such as food insecurity and housing instability, are known to contribute to poor health outcomes and inequities. Our ability to study these linkages is limited because SDoH information is more frequently documented in free-text clinical notes than structured data fields. To overcome this challenge, there is a growing push to develop techniques for automated extraction of SDoH. In this study, we explored natural language processing (NLP) and inference (NLI) methods to extract SDoH information from clinical notes of patients with chronic low back pain (cLBP), to enhance future analyses of the associations between SDoH and low back pain outcomes and disparities.

**Methods:** Clinical notes (n=1,576) for patients with cLBP (n=386) were annotated for seven SDoH domains: housing, food, transportation, finances, insurance coverage, marital and partnership status, and other social support, resulting in 626 notes with at least one annotated entity for 364 patients. We additionally labelled pain scores, depression, and anxiety. We used a two-tier taxonomy with these 10 first-level ontological classes and 68 second-level ontological classes. We developed and validated extraction systems based on both rule-based and machine learning approaches. As a rule-based approach, we iteratively configured a clinical Text Analysis and Knowledge Extraction System (cTAKES) system. We trained two machine learning models (based on convolutional neural network (CNN) and RoBERTa transformer), and a hybrid system combining pattern matching and bag-of-words models. Additionally, we evaluated a RoBERTa based entailment model as an alternative technique of SDoH detection in clinical texts. We used a model previously trained on general domain data without additional training on our dataset.

**Results:** Four annotators achieved high agreement (average kappa=95%, F_1_=91.20%). Annotation frequency varied significantly dependent on note type. By tuning cTAKES, we achieved a performance of F_1_=47.11% for first-level classes. For most classes, the machine learning RoBERTa-based NER model performed better (first-level F_1_=84.35%) than other models within the internal test dataset. The hybrid system on average performed slightly worse than the RoBERTa NER model (first-level F1=80.27%), matching or outperforming the former in terms of recall. Using an out-of-the-box entailment model, we detected many but not all challenging wordings missed by other models, reaching an average F_1_ of 76.04%, while matching and outperforming the tested NER models in several classes. Still, the entailment model may be sensitive to hypothesis wording and may require further fine tuning.

**Conclusion:** This study developed a corpus of annotated clinical notes covering a broad spectrum of SDoH classes. This corpus provides a basis for training machine learning models and serves as a benchmark for predictive models for named entity recognition for SDoH and knowledge extraction from clinical texts.

## Introduction

Adverse social determinants of health (SDoH), or social risk factors, such as food insecurity and housing instability, are recognized for their deleterious impacts on health outcomes and disparities^1^. There is growing recognition of the role of social risks in chronic low back pain (cLBP), as highlighted in a recent systematic review that found strong associations of cLBP prevalence with educational attainment and socioeconomic status^9^. Outcomes for cLBP, a leading cause of disability worldwide^3–5^, are known to be worse in patients who are economically and socially disadvantaged^6–10^. This can be attributed in part to treatment biases, including greater provision of non-evidence-based care^9,11,12^, as well as patients’ prior experience of discrimination^6,13^ and beliefs about pain and pain treatment^9,14^, which may influence engagement with the treatment offered. Much of the current research exploring disparities in cLBP care has been limited to stratifying analyses by socioeconomic status and race, as social risk data are not readily available in electronic health records (EHRs). While some social risk information exists in structured data fields, such as patient demographics and problem lists, these data are known to be under identified by clinical teams, under reported by patients, and under documented in structured fields^15–22^. When clinical teams screen for and identify social risks, that information is more often documented within free-text fields, or unstructured data^18,21^. Novel approaches to identifying SDoH in EHRs, such as Natural Language Processing (NLP) and other machine learning techniques, leverage existing health information technology to scan unstructured data fields, resulting in automated extraction^23^.

Several studies have applied NLP methods to obtain SDoH information from clinical notes. Methods have included regular expressions^24^, neural networks^25^, and rule-based algorithmic approaches^26^. Named entity recognition (NER), an NLP task of extracting phrases and their positions from texts, has been most frequently used. SDoH such as housing situation, finances, and social support^27^ have been less studied using NLP methods than behavioral determinants of health (BDoH), such as smoking status, and substance and alcohol use^23^. For BDoH, both rule-based and machine learning approaches to NER have been applied^23^. SDoH, most notably housing situation, have most commonly been identified through rule based approaches such as keyword matching^23^. Most other SDoH domains, such as transportation access and finances, have been understudied by either technique. There has been no comprehensive comparison of the strengths and weaknesses of ML and rule-based methods when applied to SDoH on various dataset sizes. We are not aware of any studies that apply NLP for SDoH for patients with cLBP.

Both machine learning (ML) and traditional rule-based system are being used to extract clinical and SDoH concepts from healthcare narratives. Tried-and-true rule-based systems such as clinical Text Analysis and Knowledge Extraction System (cTAKES)^28^, leveraging a Unified Medical Language System (UMLS) ontology developed by National Library of Medicine (NLM), could provide reasonable performance with no training data by leveraging standard ontologies and rule-based pattern matching^29,30^. Modern machine learning techniques have revolutionized NLP and natural language understanding, with self-supervised methods such as BERT^32^, allowing for efficient utilization of large amounts of un-labelled data. For optimal performance in supervised tasks in specialized domains, however, these techniques may require corpora of several (tens of) thousands of documents of manually annotated text^33^. While still providing important utility in domains where labelled data is scarce^34^ and being more interpretable than ML, rule-based systems often yield to ML methods in performance^35,36^. In terms of development costs, both rule-based and machine learning based models are resource intensive. For rule-based models, significant labor is required for manual vocabulary tuning. On the other hand, machine learning models require manually annotated data. Still, improvement of machine learning systems is normally less demanding in terms of technical development effort as compared to rule-based systems.

While several applications of natural language inference (NLI) methods, such as Question Answering (QA)^37–39^ and Recognizing Textual Entailment (RTE)^40–42^ have been pursued in the wider medical domain, we were not able to find any SDoH or BDoH studies leveraging these new methods. The fact that this has not been attempted is surprising, as one would expect NLI models to generalize to SDoH and BDoH better than other clinical domains. Knowledge about social and behavioral clues may be more widely available than knowledge about anatomy and physiology, which is necessary for understanding clinical specialties, in general domain texts (i.e., fiction and news) that state-of-the-art NLI models are predominantly trained on. This prompted us to apply NLI models in our study. Out of NLI approaches, we chose to use entailment (RTE) models instead of QA models^41^ given our interest in off-line knowledge extraction (i.e., without a need to present to a clinician for immediate decision making) with a pre-defined ontology in the setting of off-line knowledge extraction (i.e., without a need to present to a clinician for immediate decision making) with a pre-defined ontology. While extractive QA models (returning a piece of original text) may not be able to summarize text into a limited set of categories by design, abstractive QA models suffer from biases of characteristic to all generative models such as ability to learn toxic and biased attitudes^42,43^ as well as exhibit learned attitudes characteristic to mental illness and addiction^44^.

In this study, our aim was to create a dataset to evaluate and compare NLP tools to extract individual SDoH from free-text clinical notes for patients with cLBP. In this study, we make several contributions to this important topic: 1) we label and evaluate a dataset with social and behavioral determinants of health in cLBP patients, 2) we tune and evaluate a rule-based NER system (cTAKES), 3) we train and evaluate two machine learning NER pipelines, and 4) we evaluate a previously trained common domain natural language inference (NLI) entailment model. We believe this study is a step forward in both applying cutting-edge NLP technology to this important topic and in improving the structured ontology for identifying SDoH and BDoH.

## Methods

### Study population

Our study population consisted of cLBP patients, defined by low back pain lasting at least three months^43^, from a large, urban academic medical center at the University of California, San Francisco (UCSF) and was approved by institutional review (IRB #19-29016). All patients were from the UCSF Integrated Spine Service (ISS), comprised of an integrated care team of psychiatrists, pain management specialists, primary care physicians, and physical therapists^44^. For all patients, we extracted progress notes, history and physical (H&P) notes, emergency department (ED) provider notes, patient instructions, and telephone encounters (TE) from the UCSF clinical data warehouse between March 2017 to April 2020. The cohort demographics is shown in Supplementary Table 1.

### Annotation ontology

To create a model for extracting named entities related to SDoH from clinical free-text notes, we defined an ontology, annotated the dataset, and tuned and trained algorithms for named entity recognition (Fig 1). SDoH domains of interest included: *Housing, Food Transportation, Finances, Insurance Coverage, Marital and Partnership Status* and *Social Support*. These SDoH domains were selected as they are commonly included in social risk screening tools that are used in clinical practice^45–49^. Additionally, we included mental health factors (anxiety and depression) as well as pain scores, as these are relevant for our ongoing work in characterizing cLBP population^2,50–54^. During the study period, none of these SDoH were systematically screened for in our healthcare system. Our ontology builds upon a selection of social risk factors identified earlier by the UCSF Social Interventions Research and Evaluation Network (SIREN)^31^ mapped to SNOMED ontology. Initial ontology containing 68 second-level classes within 10 first-level classes was pruned after the label review step, as described in the next section, down to 52 second-level classes. The final ontology is shown in Fig 1B and Supplementary Table 2. Entities within the first-level classes not captured by specific second-level labels due to ambiguity or where low occurrence did not warrant a separate group were designated by a second-level label of “NA” for “not applicable”.

**Figure 1.**
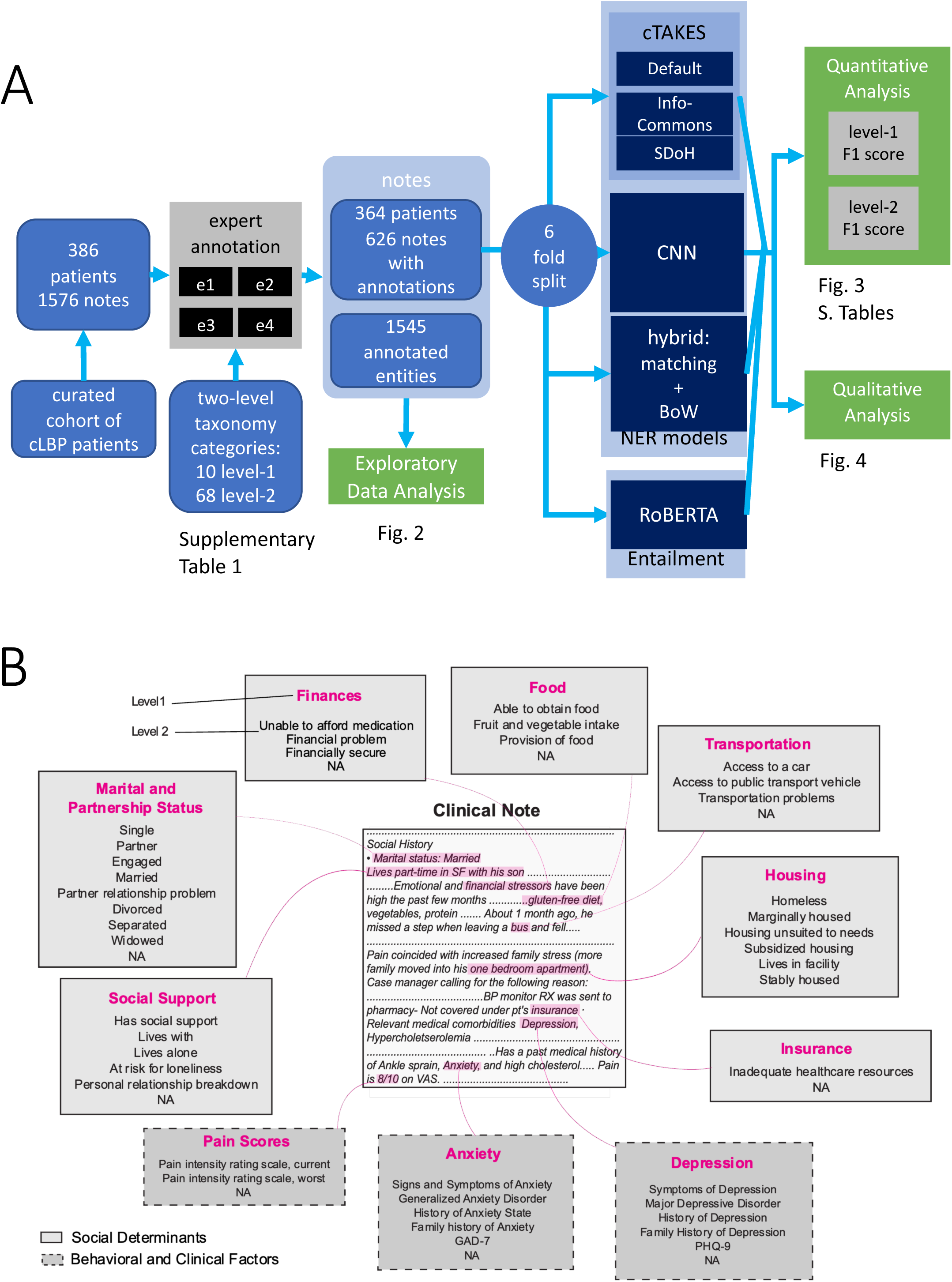
Study design. A. Workflow of the study. B. Annotation ontology. Clinical notes were annotated such that text relevant to the seven studied social risk factors (solid border) or three clinical factors (dashed border) were marked. Two levels of labels were used, such that the second level was a subcategory of the first. Level 2 labels for each level 1 annotation are shown in descending order of frequency. Level 2 annotations that comprised less than 1% of the group’s annotations are not shown. Text that can be classified to the first level but not the second due to ambiguity or low frequency is designated as “NA”. Examples of selected text are shown within the hypothetical clinical note.

### Manual Annotations

The notes were annotated according to a pre-defined two-level taxonomic ontology by four trained annotators using MAE labelling software^55^ for NER task. That is, words or phrases describing a relevant class were highlighted (as shown in Fig 1B and 4). Each note was annotated by at least two annotators (Supplementary Figure 1A). Notes were additionally spot-checked and corrected. Initial annotations occasionally contained overlapping or duplicate labels that were subsequently resolved using Python spacy package. Heuristics for duplicate resolution aimed to both maximize span length for each resulting label and to conform semantics (e.g., “lives at home with her boyfriend” may be labelled with overlapping labels describing Housing, Social Support, and Marital and Partnership Status; the corrected version would contain “lives at home” for Housing, “with her” for Social Support, and “boyfriend” for Marital and Partnership Status). Labels were further manually reviewed and rare second-level categories with low agreement and consistency labels from initial ontology were combined with more general second-level categories after data inspection. E.g., *Depressed mood* and *Symptoms of depression* were aggregated under the latter. Thus 68 initial second-level classes were pruned down to 52. After all pre-processing steps, document-level inter-rater agreement was evaluated using span-level F_1_ and document-level Cohen’s kappa^56^:

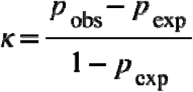

Where *p*_obs_ is the fraction of observations where experts agreed and *p*_exp_ is the expected probability of agreement between the two experts at random. For document-level evaluation, the presence or absence of at least one annotation of a given class in a clinical note was considered a binary event. Presence or absence was compared between each pair of annotators in a set of notes that they had both reviewed. A kappa per annotator pair was calculated by weighting each category with its mean occurrence according to either annotator (as described in Supplementary Methods). The final kappa was calculated as an average weighted by the number of notes annotated by each pair of annotators. We chose F_1_ as a span-level metric as it is both symmetric (as kappa is) and is less sensitive to number of true negatives, which are abundant and would dominate in NER tasks. In downstream applications, such as training and model evaluation, annotations from each expert were included. For subsequent analysis, the data was split in 6 folds, while stratifying by the number of entities and number of annotators per note and keeping multiple annotations of the same note in the same fold.

The variance of annotation frequencies was analyzed using ANOVA in R software. Pairwise difference between models was assessed using Wilcoxon test, based on F_1_ scores from each 6-fold cross-validation split.

### cTAKES configuration

We used three configurations of cTAKES software^28^. cTAKES is a modular framework developed for named entity extraction from medical texts that leverages UMLS vocabulary^57^. We used (1) a basic default out-of-the box configuration, (2) an InfoCommons configuration^58^ developed for general purpose medical texts, and (3) a customized configuration that was tuned to identify pain scores, mental health outcomes (anxiety and depression), and our SDoH domains. Tuning was done using an unlabeled set of notes and the labeling ontology definition. See Supplementary Methods for a detailed description of these three configurations.

### Spacy-based CNN model configuration

Machine learning for named entity recognition (NER) was performed with Spacy software^59^ (https://spacy.io/) using a transition-based parser leveraging a transition-based parser based on either convolutional neural network (CNN)^60^ or RoBERTa^61^ model, targeting the NER ontology described above. Word embeddings were initialized with the values from the “en_core_web_md” spacy model. The model was trained with a variable batch size of 100 to 1000 examples, drop-out of 10%, learning rate of 0.0002, and for maximum 30 epochs with early stopping. Model was trained and evaluated in nested 6-fold cross-validation, where four parts of data was used as training set, one part as validation set, and one part as test set.

### Hybrid model leveraging pattern matching and classification with Bag-of-Words

A classification model was applied to text extracted based on keyword matching patterns, which were compiled for each first-level class based on subject matter identification and literature. A JSON file of the matcher patterns is included in the code repository (https://github.com/BCHSI/social-deternimants-of-health-clbp). Text was extracted around the matching pattern (±4 tokens), pre-processed by lemmatizing and removing stop words, then embedded using a term frequency-inverse document frequency (TF-IDF) vectorizer. Extracted text for each of the first-level set of patterns was then classified using a Bag-of-Words (BoW)^62^ multinomial logistic regression model into respective second-level subclasses according to manual labels, with unmatched text spans receiving a special exclusion label. Model was trained and evaluated in 6-fold cross-validation as described for CNN model above.

### Evaluation of NER models

The results of the automated extraction systems were evaluated using precision, recall, and F1 scores. The metrics were calculated on two levels of the labeling taxonomy (first-level and second-level). For cTAKES model, predicted UMLS terms overlapping ground truth labels were manually annotated as being synonyms, children, or wrong matches of respective first-level and second-level ground truth categories, while accounting for negation, history, and family history modifiers. Synonyms and child concepts were subsequently scored as matches. The scores were calculated in using scikit learn package, with partial overlaps counted as matches. Pairwise comparison between model performance metrics was performed using paired t-test across all cross-validation folds (for CNN and hybrid models) or respective data split folds (for cTAKES), Variation of model performance was analyzed using ANOVA and nested linear model ANOVA.

### Evaluation of an entailment model

A RoBERTA-based^61^ entailment model, previously fine-tuned in an adversarial human-in-the loop setting^63^, was evaluated without additional training on our data. Given two sentences, the entailment model produces probabilities premise and hypothesis for three categories: entailment (premise statement is consistent with hypothesis), contradiction (premise not consistent with hypothesis), or neutral relation (premise indicates neither consistency nor contradiction with hypothesis)^40^.To evaluate performance of entailment model, a set of hypothesis sentences was constructed based on NER labels. E.g., “Has access to car” category was phrased as “Patient has access to a car”, “Patient drives a car”, and “Patient drives a vehicle”. Classes with final node labelled as “NA” were phrased as “This describes patient’s (first-level topic)” and were excluded from precision analysis as more specific categories would fall under that premise (See Supplementary Table 3, column “premise”). The word “patient” was replaced with a gender-specific pronoun for quantitative evaluation. Qualitative comparison of using “patient” vs gender specific pronouns indicated that replacement increases accuracy (Supplementary Methods). Each premise sentences (i.e., original text containing a label) was fed together with the each of the hypothesis sentences (See example in Fig 4). For computation expediency, only sentences with at least one NER label were examined. The results were aggregated per premise sentence (i.e., original text) and label combination either (1) by taking an average rate of entailment prediction over alternative hypotheses (“mean”) or (2) by scoring a match if at least one of the hypotheses was deemed to be entailed (“max”). Data was split into 6 folds matching the folds of NER analysis. For classes and fold sets, where positive prediction was missing and thus precision was undefined, F_1_ was assigned the value of recall. Pain scores, as well as PHQ-9 and GAD-7 scores were excluded from this analysis as they were originally included with the goal of integer value extraction and are not easily amenable for textual entailment task.

## Results

### Gold Standard Dataset Annotation

Four trained annotators annotated a set of 1,576 clinical notes with an average inter-rater variability of 95.28%, as measured by Cohen’s kappa (per-annotator metrics are shown in Suppl. Table 3) and weighted F_1_ of 91.20% and macro average F_1_ of 88.93% (Suppl Table 4). The inter-rater agreement per each label is shown in Suppl. Table 5. Of the 1,576 notes, 39.72% (626) contained at least one entity and was reviewed by at least two annotators (Supplementary Fig 1A). Notes without annotations were discarded from further analysis. ED notes contained on average the most annotations (5.6), followed by H&P (6.2), Progress notes (4.4), TE (1.8), and Patient Instructions (1.5). The number of annotations varied significantly with note type, but not with the annotator on aggregate (*p*=5e-14 and *p*=0.3 in ANOVA). The number of labels differed between note types (Fig 2). For example, *Marital and partnership status* and *Pain Scores* were frequently found in H&P and Progress notes, but rare in ED provider notes; *Insurance* coverage was mainly present in TE and Patient instructions notes. A detailed breakdown of variance p-value per note type and annotator is presented in Suppl Fig 1B. All categories except *Pain Scores, Depression, Finances*, and *Food* were significantly associated with the note type (*p*<0.05 in ANOVA). When annotations were present in a note, the association between the length of note and the number of annotations was significant (*p*<0.001, linear regression R^2^ = 0.52, Supplementary Fig 1C).

**Figure 2.**
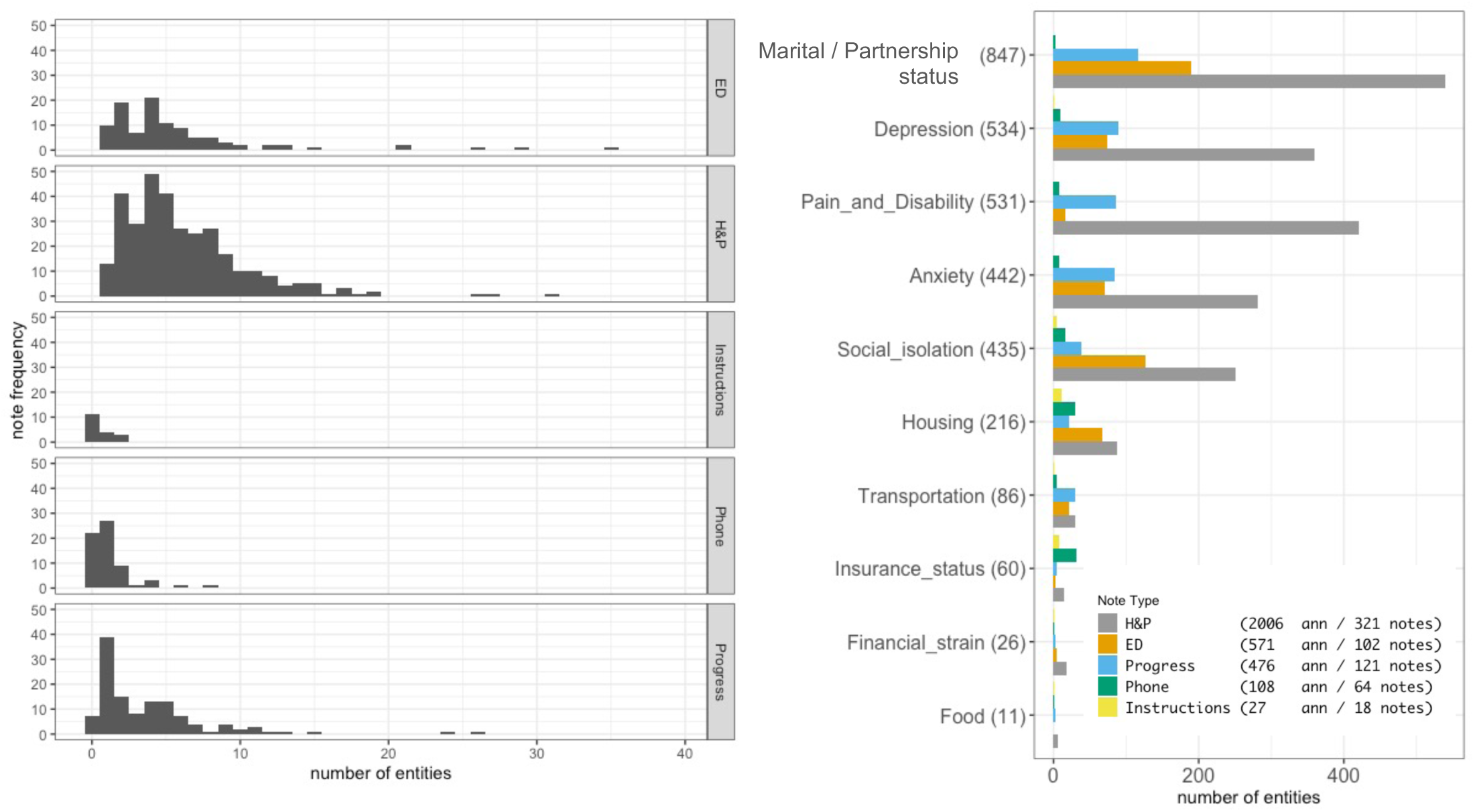
Analysis of Annotations. Note types are associated with different proportions of social domains. The total number of socially relevant spans are shown above each note type bar. History and progress notes, ED provider notes, and progress notes are more likely to have information on marital or partnership status than telephone encounter and patient instruction notes. Exploratory data analysis. A. Histogram of number of entities in different note types. B. Number of entities per note type and first-level annotated domain.

### Tuning cTAKES tool for rule-based NER

We tuned and assessed cTAKES for the task of NER of targeted SDoH domains, pain scores and anxiety and depression. As the out-of-the-box version of cTAKES has shown limitations^58^, we used a version of cTAKES configured for high throughput NER at our institution (“InfoCommons” configuration), and additionally customized cTAKES to identify SDoH for the purpose of this study (“SDoH” configuration). Tuning resulted in performance improvement across the three configurations of cTAKES (Fig. 3B, Supplementary Table 7), with weighted F_1_ achieving 38.95%, 37.95%, and 47.11% on first level for the default, InfoCommons, and SDoH versions respectively. Performance at the final (second) level of taxonomy was poor for all configurations (17.48%, 22.23%, and 34.32%, respectively). The default configuration of cTAKES captured *Depression* (F_1_ = 70.39%), *Anxiety* (F_1_ = 74.46%), and *Marital and Partnership Status* (first-level F_1_ = 45.15%) concepts relatively well (see Fig 3 and Suppl Table 6) but performed poorly on other categories (first-level F_1_<25%). Additional tuning improved the performance in *Marital and Partnership Status* (first-level F_1_=63.80% for SDoH configuration, *p*=1e-05 compared to the default configuration). However, tuning also lead to a minor degradation of performance for the categories of Anxiety (down to first-level F_1_ = 63.98%), and Depression (first-level F_1_ = 68.58%). The observed false negatives (resulting in suboptimal recall, 57.20% at the first taxonomy level for SDoH cTAKES) were commonly due to free-text wording that may not use keywords that cTAKES is configured to look for. Often wrong *history* or *negation* modifiers were attributed to the terms due to challenges with sentence boundary segmentation. SDoH words and abbreviations are occasionally misinterpreted as abbreviations from other semantic domains (e.g., “bus” and “bf” in Fig. 4). Additional qualitative analysis is provided in Supplementary Results.

**Figure 3.**
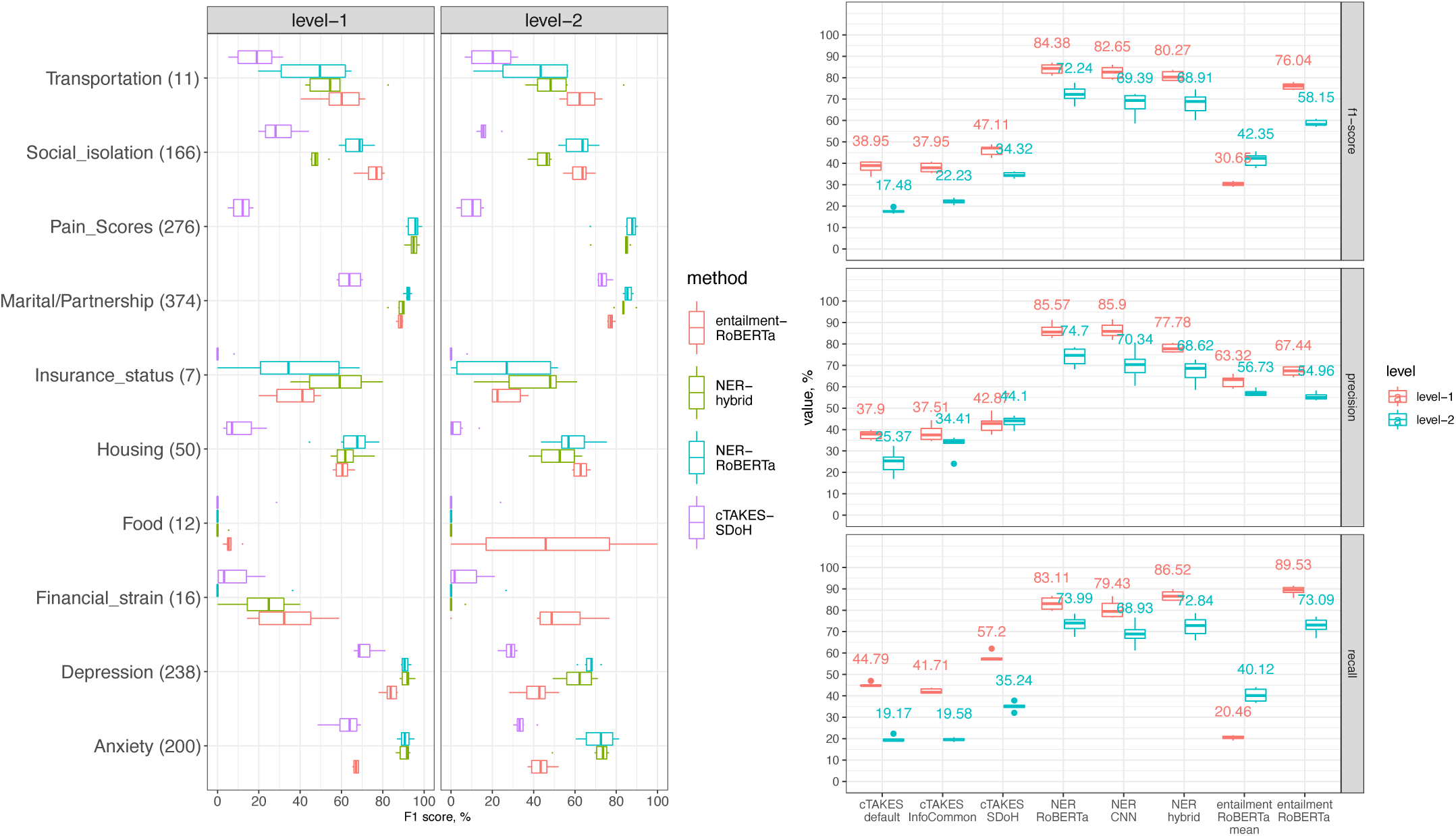
Comparison of model performance. A. Comparison of F1 performance in four best performing models per model class. Second levels metrics are aggregated using weighted average over first level domains. B. Comparison of F1, precision, and recall in all studied models. Metrics are aggregated using weighted average.

**Figure 4.**
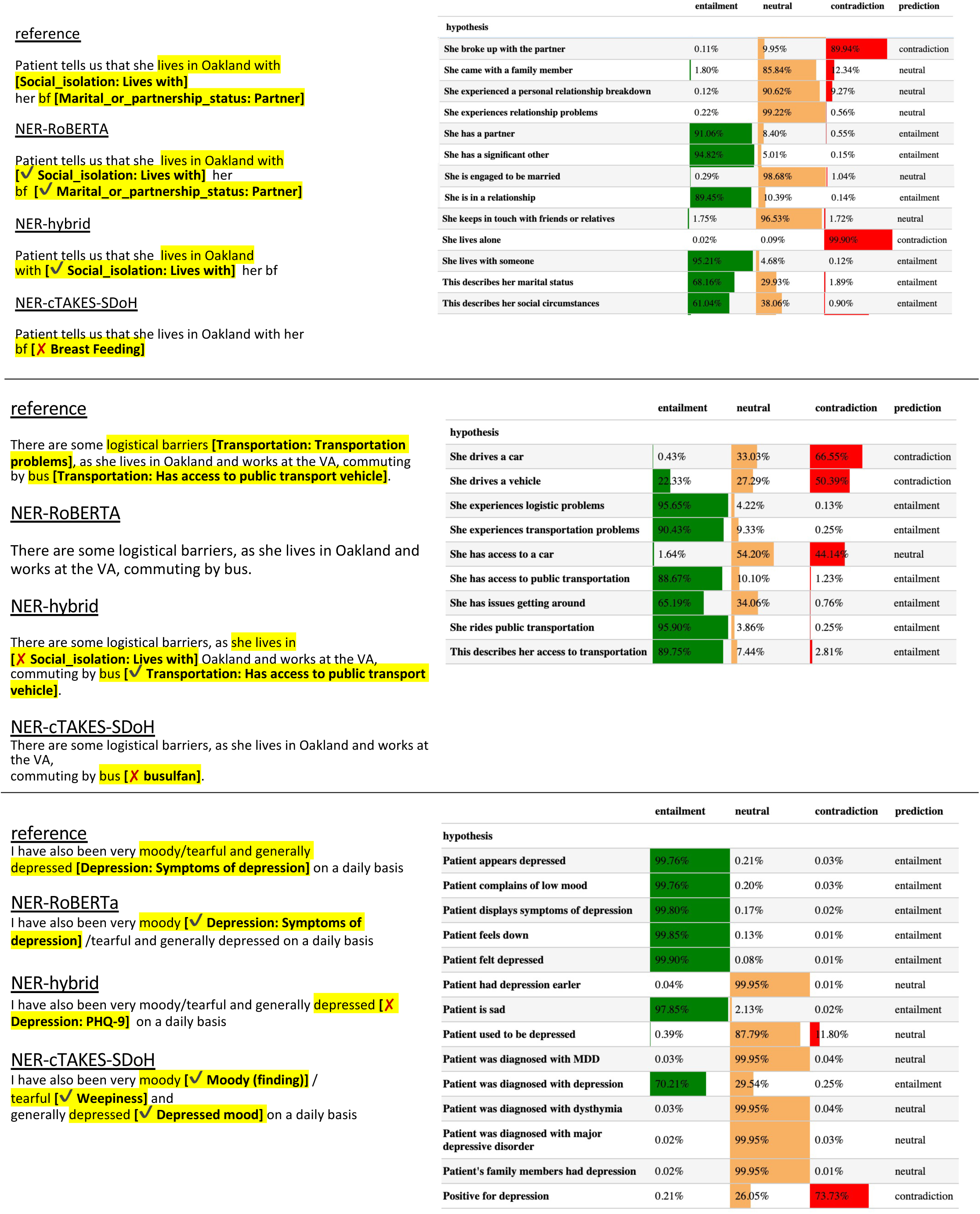
Qualitative analysis. Examples of predictions from four best models per model class. Left: NER models. Right: RoBERTA entailment model. Probabilities of three possible relations are shown as shaded horizontal bars and numerically together with a final relation prediction.

### Machine learning based NER prediction with CNN and RoBERTa

Next, we trained and evaluated performance of a CNN NER model implemented using spacy software. CNN NER and RoBERTa NER models achieved an average weighted F_1_ of 82.65% and 84.38% respectively on first-level and 69.39% and 72.24% on second-level (see Fig 3, Supplementary Tables 6 and 7). On more granular level, RoBERTa model outperformed CNN in most categories, reaching significant level only in category of Depression (second level, *p*=0.043) and approaching significant level for Social Support (*p*=0.08 and *p*=0.06 for first and second level respectively). Similar to cTAKES, some of the false positives in machine learning results were due to misinterpretation of context. For example, the word “depressed” referencing an anatomical finding, was identified as a mental health symptom. Lack of context sensitivity was a widespread cause of mislabeling of housing related concepts. For example, “plan to discharge home” does not always reflect a patient’s housing status, and “works as a home CNA” does not refer to patient housing status. In some cases, negation markers such as “denies” were not accounted for. Misspellings were also missed, such as “derpession” for “depression.”

### Hybrid text extraction and classification system

Next, we applied a hybrid approach, which extracts relevant text segments by pattern matching and classifies with a logistic regression BoW model. The hybrid model displayed a high recall (86.52% at first level), outperforming other methods (Fig 3, Supplementary Table 6, 7, and 8), with an average first-level F_1_= 80.27%. Several cases of failure may be attributed to fundamental limitations of the underlying BoW model, such as cases that refer to people other than the patient, e.g., “her father lives with a partner” was interpreted as “Lives with” category pertaining to the patient.

### Comparison of NER model performance

The machine learning and hybrid models significantly outperformed the rule-based system in terms of F_1_ on the first level (*p*<1e-5) and the second level (*p*<0.03) of the taxonomy for all classes except *Finances* and *Food* (Fig 3, Suppl. Table 6 and 8.) While the NER-RoBERTa model achieved the highest first-level F_1_=84.38% among all models (*p*<0.04), it scored on par (72.24%) with the hybrid model (68.91%) at the second level F_1_ (*p*=0.13). The hybrid model had a consistently higher recall for all first-level categories but was outperformed by the RoBERTa based NER in terms of first-level F_1_ in *Marital status (p=0*.*03)* and both first and second level in *Social isolation (p<0*.*001)*. All NER models performed poorly for the classes of *Food* and second level domains with low F_1_ agreement (35%--60%, Suppl. Table 5), performed poorly in all methods (Suppl. Fig 2), including classes of *Depression: NA, Marital or Partnership status: NA, Social Support: NA, Housing: Housing unsuited to needs, Marginally housed*. To evaluate contribution of various factors to the model performance, we conducted a multifactor ANOVA of F_1_ metric. Both the method and the ontology label are significantly associated with the performance, but not data split fold (*p*<2e-31 Suppl. Table 9). Additionally, per-class performance was correlated across methods (*p*<2e-5, Suppl. Table 10). Inter-rater agreement F_1_ explains 6.76% of between-label variation in F_1_ model performance (*p*=2e-20 in one-way ANOVA).

### Evaluation of transformer-based models for entailment

We hypothesized that general domain NLI models may be directly applied to SDoH and BDoH domains due to semantic overlap between SDoH and general domain texts. Thus, we evaluated performance of an entailment model^63^ without fine-tuning, whereby a subset of premise sentences that contained labelled named entities was taken from clinical notes and was passed together with a set of hypothesis sentences compiled to reflect semantics of the respective named entities. The aggregated metrics are shown in Fig. 3 and examples of prediction are shown in Fig 4, while granular metrics are presented in Supplementary Fig 2. Max-aggregation (relying on an assumption that at least one of the hypotheses per class must holds) performed better than mean-aggregation (one assuming all hypotheses must hold), F_1_=76.04% vs F_1_=30.65% at the first level respectively. The model scored best on categories of *Marital Status, Depression, Anxiety*, and *Social Support* (F_1_>70%). The entailment model outperformed the NER models evaluated above in the categories of *Transportation, Finances*, and *Food* (Fig. 3). Qualitative analysis reveals sensitivity of the results to hypothesis wording, which in turn may be sensitive to the way the patient is referred to for proper reference resolution. Additionally, entailment models performed poorly on long lists such as lists of family history conditions and lists of diagnosis due to both sentence segmentation issues and difficulties with incomplete sentences. Further qualitative details are provided in Supplementary Results.

## Discussion

In this study, we developed a corpus of annotated clinical notes covering a broad spectrum of SDoH domains, together with anxiety, depression, and pain scores for cLBP patients. This corpus provides a basis for training machine learning models and serves as a benchmark for both machine learning and other types of predictive models for NER for SDoH. H&P notes were the richest source of SDoH, while patient instructions contained little relevant information.

By evaluating various NER pipelines, we identified strengths and weaknesses of both rule-based (cTAKES) and machine learning based approaches for identifying SDoH domains. For most of our evaluated categories, the machine learning methods outperformed cTAKES, except for *Finances* and *Food security*, domains for which very little training data was available. This may be expected as cTAKES is rule-based and thus does not require training data and relies instead on the richness of synonym vocabularies in NLM Metathesaurus. Some of cTAKES’s performance limitations appeared to be due to a failure to account for non-affirmative mentions in questionnaires or missing rare keywords. Inclusion of these keywords may improve cTAKES recall. Additionally, poor cTAKES performance on the second-level taxonomy may be attributed to difficulty detecting nuanced differences, such as distinguishing between symptoms (e.g., anxiety) vs disorders (e.g., generalized anxiety disorder), which depends heavily on the context. Polysemous abbreviations present a challenge even for human intelligibility in general medical practice^64^, and are even harder for multi-domain rule-based tools such as cTAKES that have no means of factoring in domain-specific contextual knowledge. Additional tuning of the cTAKES model based on the wordings present in annotations obtained in this study may overcome some of these challenges. Due to study resource constraints, we were limited in our ability to continue tuning cTAKES, but our work provides an understanding of the current limitations and shortcomings that may be improved upon.

Comparing all models, we noticed that the detection performance of different SDoH domains was correlated across methods, which suggests that model performance is limited by both fundamental variability of phrasing in free text notes, as well as training data size. Additional analysis showed that inter-rater variability explains this correlation to a small degree. In our dataset, the least documented categories were insurance status, transportation access, food security, and finances, which may be due to a low occurrence of these risk factors in our population; lack of screening for social risk factors; and/or under-documentation of these risks, even when they are known to the health care team. Annotations were also limited by the quality of notes, which could hinder our ability to categorize potentially relevant phrases. Most of these rare categories were detected with low performance. On the other hand, categories of higher frequency such as depression, anxiety, pain scores, as well as marital and partnership status, were the categories detected most accurately by all methods. Machine learning and hybrid methods outperformed rule-based method (cTAKES) for most classes except *Finances* and *Insurance status* which were detected best by the hybrid model. These classes exemplify data scarce situations where rule-based systems may confer advantage^34,65^, in pure form or as a component of pipeline, as they do not rely on training data. Thus, on average in terms of F_1_, CNN model performed best, yielding up to the hybrid and rule-based models for a few data-poor classes.

Evaluation of an out-of-the-box entailment RoBERTa^66^model trained on general domain texts, yielded promising results. Without fine-tuning, this model performed comparably to machine learning models trained on our corpus in most categories. Direct comparison of F_1_ metric examined in our study (76%) to the accuracy reported in the original study by Nie and colleagues^66^ (50%–93% depending on test set) is not possible, as it would require additional manual annotation to account for cases of contradiction relation. However, both qualitative and quantitative analysis suggests that the model generalizes reasonably well to SDoH domains. As performance of the out-of-the box entailment model exceeded the characteristics of widely accepted rule-based pipeline (cTAKES), with recall of the former nearing 90% without fine-tuning, we believe that entailment models maybe readily deployed in user-facing tools for clinical text exploration and retrieval, such as EMERSE^66^, thus allowing researchers to query data based on custom criteria. However, sensitivity of the model to the hypothesis wording observed in this study needs to be taken into consideration when creating fine-tuning data, directly applying to new data, or querying with different hypotheses. Additional fine-tuning on medical- and SDoH-related corpora may further increase performance and should be further explored.

Our study should be interpreted with consideration of its limitations. First, the generalizability of the models validated here may be affected by the fact that clinical notes came from an urban academic medical center, where fewer patients may experience — or fewer providers may be aware to inquire and document — social risk factors, versus the Veterans Affairs or safety-net hospitals, where much of the current SDoH NLP literature is derived. Our study may therefore be better generalizable to other urban academic medical centers than to other settings. This may be a limitation while applying models developed in this study to different kind of institutions, even when they are located in the same geographic area. Related, our study focused on notes for patients with chronic LBP and not other conditions. We included, however, a diverse set of note types from different settings that may improve the generalizability to patients with other conditions. Further validation of the presented models across different medical centers and for patients with different medical conditions is needed to understand their wider applicability. Second, as previously noted, some SDoH domains had particularly low counts of annotations, such as food security and finances. We erred on the side of not making assumptions when annotating text, as notes were not always written clearly due to grammatical errors and/or the shorthand format of notes, which led many annotations to fall under the “NA” (other) category. The strength and capacity of our models overall are limited by the quality of notes that were annotated. Third, dictionaries constructed during tuning of rule-based and hybrid model were composed based on the complete dataset (before annotation), thus, data leakage may not be completely excluded. Lastly, as noted above, our study’s resources limited our ability to continue to fine tune cTAKES and further refine our models overall. Differences in performance between the models may therefore exaggerate the limitations of cTAKES for identifying SDoH as compared to other methods. This study, however, outlines how cTAKES, as well as the other models, can be improved upon.

## Conclusion

This study is an important step toward understanding the differences between, and strengths and limitations of, a diverse set of NLP methods for detecting SDoH domains within clinical notes. To our knowledge, this is the first study to compare a broad panel of ten domains of SDoH, BDoH, and pain scores with 52 granular sub-domains. This study lays the foundation for better detecting social risk factors in chronic low back pain patients, which can advance our understanding of how social risks impact low back pain treatment access, utilization, and outcomes. Our findings and open-source methods can also be applied to other settings and patient populations, to fuel the growing momentum to apply machine learning techniques to the detection of SDoH in EHRs.

## Data Availability

Raw data analyzed in the present study are not available due to privacy concerns. Resulting summary statistics are contained in the manuscript.

https://github.com/BCHSI/social-deternimants-of-health-clbp

## Acknowledgements

This work was supported by Back Pain Consortium (BACPAC) grant, UCSF Core Center for Patient-centric Mechanistic Phenotyping in Chronic Low Back Pain (UCSF REACH), UCSF Social Interventions Research and Evaluation Network (SIREN), the Innovate for Health program, including the UC Berkeley Institute for Data Science, the UCSF Bakar Computational Health Sciences Institute and Johnson & Johnson (to BL). We would like to acknowledge Madhumita Sushil and Hunter Mills for their advice and comments on this manuscript, and Sara Jan, Matthew Lee, and Rafael Lee for their work as annotators.

## Supplementary Figures

**Supplementary Fig 1.**
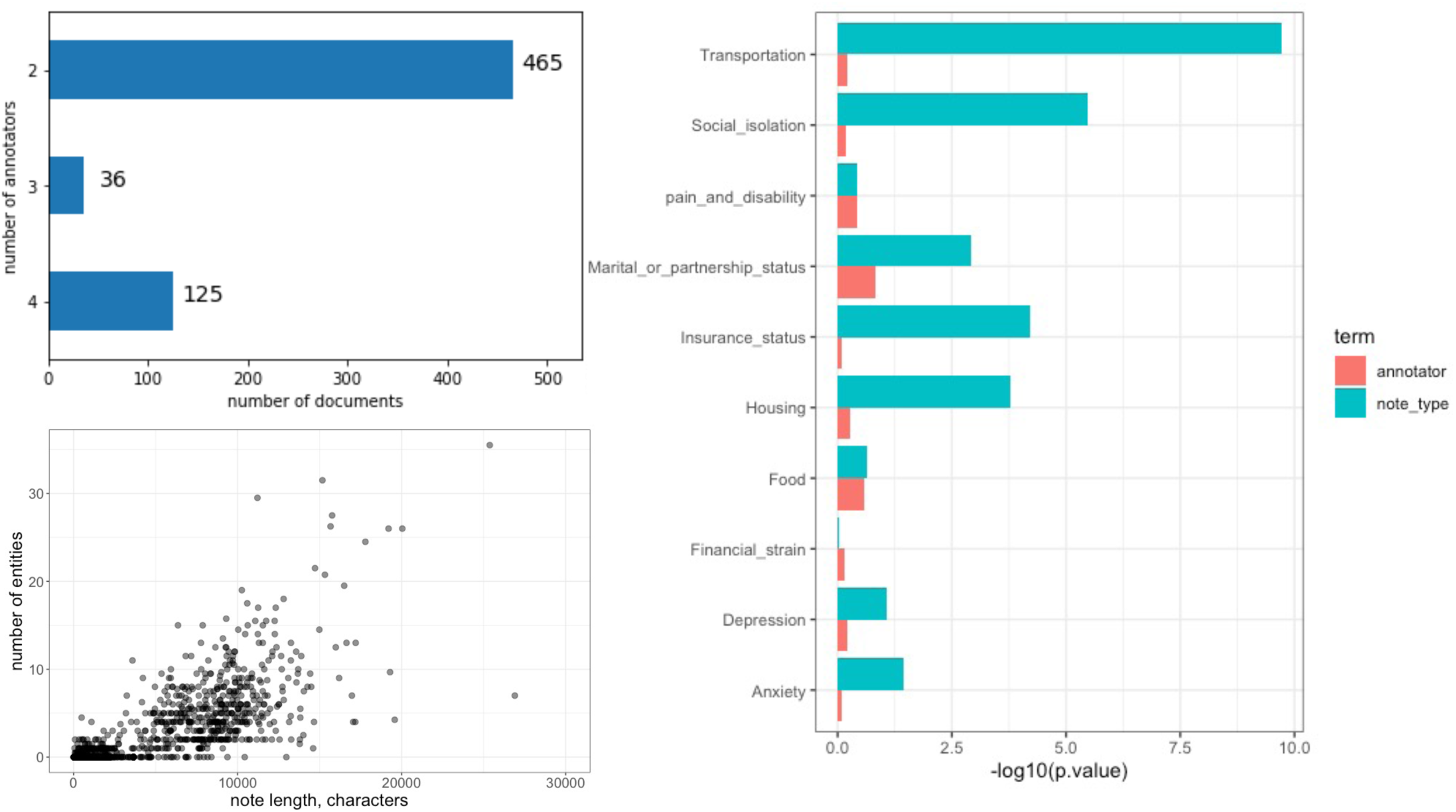
Annotations A. Number of annotators per document B. ANOVA of frequency of annotations note per annotator and note type C. Scatter plot of number of entities vs note length

**Supplementary Figure 2.**
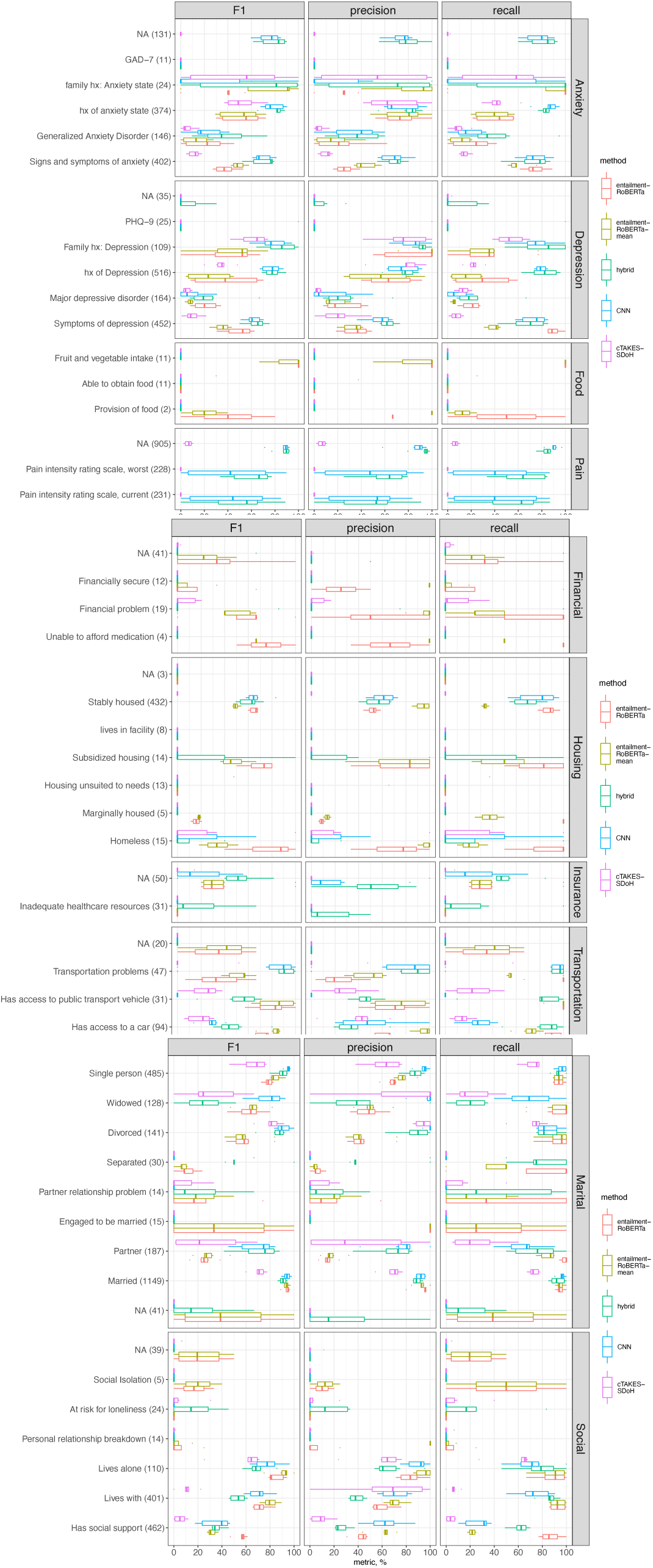
Granular Level-2 metrics

## Notes

### Competing Interest Statement

DL is a shareholder of Crosscope Inc and SynthezAI Corp.
BL is supported by Innovate for Health Data Science Fellowship from Johnson & Johnson.
PLA received funding from REAC RAP UCSF through UCSF.
EDM received support from Hellman Fellows Fund Payment, Episcopal Health Foundation, and REAC RAP UCSF through UCSF.
SP received support from Back Pain Consortium (BACPAC) grant through UCSF

### Funding Statement

This study was supported by Back Pain Consortium (BACPAC) grant, UCSF Core Center for Patient-centric Mechanistic Phenotyping in Chronic Low Back Pain (UCSF REACH), UCSF Social Interventions Research and Evaluation Network (SIREN), the Innovate for Health program, including the UC Berkeley Institute for Data Science, the UCSF Bakar Computational Health Sciences Institute and Johnson & Johnson (to BL).

### Author Declarations

IRB of University of California, San Francisco gave ethical approval for this work

## References

1. Hatef, E. et al. Integrating social and behavioral determinants of health into patient care and population health at Veterans Health Administration: a conceptual framework and an assessment of available individual and population level data sources and evidence-based measurements. AIMS Public Health 6, 209–224 (2019).

2. Karran, E. L., Grant, A. R. & Moseley, G. L. Low back pain and the social determinants of health: a systematic review and narrative synthesis. PAIN 161, 2476–2493 (2020).

3. James, S. L. et al. Global, regional, and national incidence, prevalence, and years lived with disability for 354 diseases and injuries for 195 countries and territories, 1990–2017: a systematic analysis for the Global Burden of Disease Study 2017. The Lancet 392, 1789–1858 (2018).

4. U. S. Burden of Disease Collaborators et al. The State of US Health, 1990-2016: Burden of Diseases, Injuries, and Risk Factors Among US States. JAMA 319, 1444–1472 (2018).

5. Dutmer, A. L. et al. Personal and Societal Impact of Low Back Pain: The Groningen Spine Cohort. Spine Phila Pa 1976 44, E1443–E1451 (2019).

6. Trost, Z. et al. Examining Injustice Appraisals in a Racially Diverse Sample of Individuals With Chronic Low Back Pain. J Pain 20, 83–96 (2019).

7. Chen, Y. et al. Trajectories and predictors of the long-term course of low back pain: cohort study with 5-year follow-up. Pain 159, 252–260 (2018).

8. Batley, S. et al. The association between psychological and social factors and spinal pain in adolescents. Eur J Pediatr 178, 275–286 (2019).

9. Anderson, K. O., Green, C. R. & Payne, R. Racial and ethnic disparities in pain: causes and consequences of unequal care. J Pain 10, 1187–204 (2009).

10. Green, C. R. et al. The Unequal Burden of Pain: Confronting Racial and Ethnic Disparities in Pain. Pain Med 4, 277–294 (2003).

11. Tait, R. C., Chibnall, J. T., Andresen, E. M. & Hadler, N. M. Management of occupational back injuries: differences among African Americans and Caucasians. Pain 112, 389–96 (2004).

12. Gebauer, S., Salas, J. & Scherrer, J. F. Neighborhood Socioeconomic Status and Receipt of Opioid Medication for New Back Pain Diagnosis. J Am Board Fam Med 30, 775–783 (2017).

13. Ziadni, M. S. et al. Injustice Appraisal, but not Pain Catastrophizing, Mediates the Relationship Between Perceived Ethnic Discrimination and Depression and Disability in Low Back Pain. J Pain (2019) doi:10.1016/j.jpain.2019.09.007.

14. Suman, A., Bostick, G. P., Schaafsma, F. G., Anema, J. R. & Gross, D. P. Associations between measures of socio-economic status, beliefs about back pain, and exposure to a mass media campaign to improve back beliefs. BMC Public Health 17, 504 (2017).

15. Vest, J. R., Wu, W. & Mendonca, E. A. Sensitivity and Specificity of Real-World Social Factor Screening Approaches. J. Med. Syst. 45, 111 (2021).

16. Hong, Y.-R., Turner, K., Nguyen, O. T., Alishahi Tabriz, A. & Revere, L. Social Determinants of Health and After-Hours Electronic Health Record Documentation: A National Survey of US Physicians. Popul. Health Manag. (2021) doi:10.1089/pop.2021.0212.

17. Wang, M., Pantell, M. S., Gottlieb, L. M. & Adler-Milstein, J. Documentation and review of social determinants of health data in the EHR: measures and associated insights. J. Am. Med. Inform. Assoc. ocab194 (2021) doi:10.1093/jamia/ocab194.

18. Hatef, E. et al. Assessing the Availability of Data on Social and Behavioral Determinants in Structured and Unstructured Electronic Health Records: A Retrospective Analysis of a Multilevel Health Care System. JMIR Med. Inform. 7, e13802 (2019).

19. Arons, A., DeSilvey, S., Fichtenberg, C. & Gottlieb, L. Documenting social determinants of health-related clinical activities using standardized medical vocabularies. JAMIA Open 2, 81–88 (2019).

20. Cottrell, E. K. et al. Variation in Electronic Health Record Documentation of Social Determinants of Health Across a National Network of Community Health Centers. Am. J. Prev. Med. 57, S65–S73 (2019).

21. Beck, A. F., Klein, M. D. & Kahn, R. S. Identifying Social Risk via a Clinical Social History Embedded in the Electronic Health Record. Clin. Pediatr. (Phila.) 51, 972–977 (2012).

22. Torres, J. M. et al. ICD Social Codes: An Underutilized Resource for Tracking Social Needs. Med. Care 55, 810–816 (2017).

23. Patra, B. G. et al. Extracting social determinants of health from electronic health records using natural language processing: a systematic review. J. Am. Med. Inform. Assoc. (2021) doi:10.1093/jamia/ocab170.

24. Chen, E. S., Carter, E. W., Sarkar, I. N., Winden, T. J. & Melton, G. B. Examining the Use, Contents, and Quality of Free-Text Tobacco Use Documentation in the Electronic Health Record. AMIA. Annu. Symp. Proc. 2014, 366–374 (2014).

25. Bejan, C. A. et al. Mining 100 million notes to find homelessness and adverse childhood experiences: 2 case studies of rare and severe social determinants of health in electronic health records. J. Am. Med. Inform. Assoc. 25, 61–71 (2018).

26. Conway, M. et al. Moonstone: a novel natural language processing system for inferring social risk from clinical narratives. J. Biomed. Semant. 10, 6 (2019).

27. Stemerman, R. et al. Identification of social determinants of health using multi-label classification of electronic health record clinical notes. JAMIA Open 4, (2021).

28. Savova, G. K. et al. Mayo clinical Text Analysis and Knowledge Extraction System (cTAKES): architecture, component evaluation and applications. J. Am. Med. Inform. Assoc. JAMIA 17, 507–513 (2010).

29. Afshar, M. et al. Natural language processing and machine learning to identify alcohol misuse from the electronic health record in trauma patients: development and internal validation. J. Am. Med. Inform. Assoc. JAMIA 26, 254–261 (2019).

30. Shoenbill, K. et al. Natural language processing of lifestyle modification documentation. Health Informatics J. 26, 388–405 (2020).

31. Arons, A., DeSilvey, S., Fichtenberg, C. & Gottlieb, L. M. Compendium of Medical Terminology Codes for Social Risk Factors. University of California, San Francisco; Social Interventions Research and Evaluation Network. (2019).

32. Devlin, J., Chang, M.-W., Lee, K. & Toutanova, K. BERT: Pre-training of Deep Bidirectional Transformers for Language Understanding. ArXiv181004805 Cs (2019).

33. Rasmy, L., Xiang, Y., Xie, Z., Tao, C. & Zhi, D. Med-BERT: pretrained contextualized embeddings on large-scale structured electronic health records for disease prediction. Npj Digit. Med. 4, 86 (2021).

34. Chiticariu, L., Li, Y. & Reiss, F. R. Rule-Based Information Extraction is Dead! Long Live Rule-Based Information Extraction Systems! in Proceedings of the 2013 Conference on Empirical Methods in Natural Language Processing 827–832 (Association for Computational Linguistics, 2013).

35. Jorge, A. et al. Identifying lupus patients in electronic health records: Development and validation of machine learning algorithms and application of rule-based algorithms. Semin. Arthritis Rheum. 49, 84–90 (2019).

36. Topaz, M. et al. Mining fall-related information in clinical notes: Comparison of rule-based and novel word embedding-based machine learning approaches. J. Biomed. Inform. 90, 103103 (2019).

37. Cairns, B. L. et al. The MiPACQ clinical question answering system. AMIA Annu. Symp. Proc. AMIA Symp. 2011, 171–180 (2011).

38. Pampari, A., Raghavan, P., Liang, J. & Peng, J. emrQA: A Large Corpus for Question Answering on Electronic Medical Records. in Proceedings of the 2018 Conference on Empirical Methods in Natural Language Processing 2357–2368 (Association for Computational Linguistics, 2018). doi:10.18653/v1/D18-1258.

39. Patrick, J. & Li, M. An ontology for clinical questions about the contents of patient notes. J. Biomed. Inform. 45, 292–306 (2012).

40. Dagan, I., Roth, D., Sammons, M. & Zanzotto, F. M. Recognizing Textual Entailment: Models and Applications. Synth. Lect. Hum. Lang. Technol. 6, 1–220 (2013).

41. Ben Abacha, A. & Demner-Fushman, D. A question-entailment approach to question answering. BMC Bioinformatics 20, 511 (2019).

42. Shivade, C. et al. Textual inference for eligibility criteria resolution in clinical trials. J. Biomed. Inform. 58 Suppl, S211–S218 (2015).

43. Deyo, R. A. et al. Report of the NIH Task Force on Research Standards for Chronic Low Back Pain. Phys. Ther. 95, e1–e18 (2015).

44. O’Neill, C. & Zheng, P. Integrated Spine Service: Putting Value into Back Pain Care. SPINELINE vol. 20 12–14 (2019).

45. Medicine, I. of. Capturing Social and Behavioral Domains in Electronic Health Records: Phase 1. (The National Academies Press, 2014). doi:10.17226/18709.

46. Medicine, I. of. Capturing Social and Behavioral Domains and Measures in Electronic Health Records: Phase 2. (The National Academies Press, 2014). doi:10.17226/18951.

47. Hager, E. R. et al. Development and Validity of a 2-Item Screen to Identify Families at Risk for Food Insecurity. Pediatrics 126, e26–e32 (2010).

48. PRAPARE_One_Pager_Sept_2016.pdf.

49. Social Needs Screening Tool Comparison Table | SIREN. https://sirenetwork.ucsf.edu/tools-resources/resources/screening-tools-comparison.

50. Pinheiro, M. B. et al. Symptoms of depression as a prognostic factor for low back pain: a systematic review. Spine J. 16, 105–116 (2016).

51. Froud, R. et al. A systematic review and meta-synthesis of the impact of low back pain on people’s lives. BMC Musculoskelet. Disord. 15, 50 (2014).

52. Hong, J. H., Kim, H. D., Shin, H. H. & Huh, B. Assessment of depression, anxiety, sleep disturbance, and quality of life in patients with chronic low back pain in Korea. Korean J. Anesthesiol. 66, 444–450 (2014).

53. Tsuji, T., Matsudaira, K., Sato, H. & Vietri, J. The impact of depression among chronic low back pain patients in Japan. BMC Musculoskelet. Disord. 17, 447 (2016).

54. Pincus, T., Burton, A. K., Vogel, S. & Field, A. P. A Systematic Review of Psychological Factors as Predictors of Chronicity/Disability in Prospective Cohorts of Low Back Pain. Spine 27, E109 (2002).

55. Rim, K. MAE2: Portable Annotation Tool for General Natural Language Use. Proc. 12th Jt. ACL-ISO Workshop Interoper. Semantic Annot. Portorož Slov. May 28 2016 6 (2016).

56. Cohen, J. A Coefficient of Agreement for Nominal Scales. Educ. Psychol. Meas. 20, 37–46 (1960).

57. Unified Medical Language System (UMLS). https://www.nlm.nih.gov/research/umls/index.html.

58. Abramowitsch, P. Apache cTAKES High Throughput Orchestration. (2020).

59. Honnibal, M. & Montani, I. spaCy 2: Natural language understanding with Bloom embeddings, convolutional neural networks and incremental parsing. (2017).

60. Honnibal, M. & Johnson, M. An Improved Non-monotonic Transition System for Dependency Parsing. in Proceedings of the 2015 Conference on Empirical Methods in Natural Language Processing 1373–1378 (Association for Computational Linguistics, 2015). doi:10.18653/v1/D15-1162.

61. Liu, Y. et al. RoBERTa: A Robustly Optimized BERT Pretraining Approach. ArXiv190711692 Cs (2019).

62. Harris, Z. S. Distributional Structure. WORD 10, 146–162 (1954).

63. Nie, Y. et al. Adversarial NLI: A New Benchmark for Natural Language Understanding. in Proceedings of the 58th Annual Meeting of the Association for Computational Linguistics (Association for Computational Linguistics, 2020).

64. Neil M Davis. Medical abbreviations with multiple meanings: A prescription for disaster. 29,.

65. Gorinski, P. J. et al. Named Entity Recognition for Electronic Health Records: A Comparison of Rule-based and Machine Learning Approaches. ArXiv190303985 Cs (2019).

66. Hanauer, D. A., Mei, Q., Law, J., Khanna, R. & Zheng, K. Supporting information retrieval from electronic health records: A report of University of Michigan’s nine-year experience in developing and using the Electronic Medical Record Search Engine (EMERSE). J. Biomed. Inform. 55, 290–300 (2015).

