## supplementary material for "Automatic Extraction of Social Determinants of Health from Medical Notes of Chronic Lower Back Pain Patients"

### **Supplementary Tables**

Supplementary Table 1. Demographics of patients

|  | **Overall** **(*n*=386)** |
| --- | --- |
| **Sex** |  |
| Female | 242 (62.7%) |
| Male | 144 (37.3%) |
| **Race** |  |
| American Indian or Alaska Native | 1 (0.3%) |
| Asian | 85 (22.0%) |
| Black or African American | 56 (14.5%) |
| Declined | 6 (1.6%) |
| Native Hawaiian or Other Pacific Islander | 4 (1.0%) |
| Other | 44 (11.4%) |
| Other Pacific Islander | 1 (0.3%) |
| White or Caucasian | 189 (49.0%) |
| **Ethnicity** |  |
| Declined | 8 (2.1%) |
| Hispanic or Latino | 35 (9.1%) |
| Not Hispanic or Latino | 342 (88.6%) |
| Unknown | 1 (0.3%) |
| **First STARTBACK score** |  |
| High Risk | 116 (30.1%) |
| Low Risk | 111 (28.8%) |
| Medium Risk | 159 (41.2%) |
| **Age** |  |
| Mean (SD) | 53.7 (17.5) |
| Median [Min, Max] | 54.0 [6.00, 98.0] |
| **BMI** |  |
| Mean (SD) | 26.1 (7.20) |
| Median [Min, Max] | 25.4 [0, 54.9] |
| **Area Deprivation Index** |  |
| Mean (SD) | 5.16 (6.78) |
| Median [Min, Max] | 3.03 [1.00, 85.0] |
| Unknown | 2 (0.2%) |
| **CCI score** |  |
| Mean (SD) | 0.659 (1.33) |
| Median [Min, Max] | 0 [0, 12.0] |
| Unknown | 156 (12.0%) |

Supplementary Table 2A. Annotation ontology

| first level | second level | canonical text | SNOMED_CT | CUI |
| --- | --- | --- | --- | --- |
| Anxiety | Signs and symptoms of anxiety | Signs and symptoms of anxiety |  | C0860603 |
|  | Generalized Anxiety Disorder | Generalized Anxiety Disorder | 21897009 | C0270549 |
|  | family hx: Anxiety state | Family history: Anxiety state | 160332003 | C0455385 |
|  | hx of anxiety state | History of anxiety state | 161470009 | C0455504 |
|  | GAD-7 | Generalized anxiety disorder 7 item score | 445455005 | C2919162 |
| Depression | Major depressive disorder | Major depressive disorder | 370143000 | C1269683 |
|  | Family hx | Family history: Depression | 160329001 | C0455383 |
|  | Depressed mood | Depressed mood | 366979004 | C0344315 |
|  | Hx of Depression | History of depression | 161469008 | C0455503 |
|  | Symptoms of depression | Symptoms of depression | 394924000 | C4716638 |
|  | PHQ-9 | Depression screening using Patient Health Questionnaire Nine Item score | 715252007 | C4275234 |
| Pain Scores | NA | Pain intensity rating scale | 425401001 | C1960637 |
|  | Pain intensity rating scale, current | Pain intensity rating scale, current | 425401001 | C1960637 |
|  | Pain intensity rating scale, worst | Pain intensity rating scale, worst | 425401001 | C1960637 |
| Housing | Stably housed | Housed | 414418009 | C0020056 |
|  | Homeless | Homeless | 32911000 | C0237154 |
|  | Marginally housed | Housing instability | 1156191002 | C5539303 |
|  | Subsidized housing | Family Received Low-Income or Subsidized Housing |  | C5204673 |
|  | Housing unsuited to needs | Housing unsuited to needs | 160721001 | C0578892 |
|  | Lives in facility | Assisted living facility patient | 11762561000119100 | C4510636 |
|  | NA | Residence and accommodation circumstances | 224209007 | C0557202 |
| Transportation | Has access to a car | Has access to a car | 160687009 | C0425253 |
|  | Has access to public transport vehicle | Has access to public transport vehicle | 413288008 | C1446359 |
|  | Transportation problems | Transport problems | 266934004 | C0425259 |
|  | NA | Access to transportation | 1137435002 | C3650743 |
| Finances | Financially secure | Financially secure | 224165005 | C0557160 |
|  | Financial problem | Financial problem | 160932005 | C0549106 |
|  | Unable to afford medication | Unable to afford medication | 454061000124102 | C4520569 |
|  | NA | Finding of financial circumstances | 365550006 | C1287207 |
| Social Support | NA | Social context finding | 108329005 | C1266850 |
|  | At risk for loneliness | At risk for loneliness | 129698004 | C1268628 |
|  | Has social support | Receives as much social support as wanted | 445091000124106 | C3853122 |
|  | Social Isolation | Social Isolation | 422650009 | C0037421 |
|  | Lives alone | Lives alone | 105529008 | C0439044 |
|  | Lives with | Household composition | 224130005 | C0595998 |
|  | Personal relationship breakdown | Personal relationship breakdown | 105414008 | C0524322 |
| Insurance status | Inadequate healthcare resources | Inadequate healthcare resources | 423593006 | C1828339 |
|  | NA | Finding related to health insurance issues | 419808006 | C1636784 |
| Food | Able to obtain food | Food security | 286441005 | C0564281 |
|  | Provision of food | Provision of food | 710925007 | C4039510 |
|  | Fruit and vegetable intake | Fruit and vegetable intake | 391129005 | C1271941 |
|  | NA | Nutritional finding | 300893006 | C0577601 |
| Marital or partnership status | Separated | Separated | 13184001 | C4721417 |
|  | Divorced | Divorce | 63234004 | C0086170 |
|  | Widowed | Widowed | 33553000 | C1510465 |
|  | Partner | Partner in relationship | 262043009 | C0682323 |
|  | Common law partnership | Common law partnership | 14012001 | C0009448 |
|  | Engaged to be married | Engaged to be married | 54986009 | C0425152 |
|  | Married | Married | 87915002 | C0555047 |
|  | Single person | Single person | 125681006 | C0037179 |
|  | Partner relationship problem | Partner relationship problem | 1040000000000 | C0236858 |
|  | NA | Marital status | 125680007 | C0024819 |

Supplementary Table 2B. Annotation ontology and entailment hypotheses

| first level | second level | CUI | hypothesis |
| --- | --- | --- | --- |
| Anxiety | Signs and symptoms of anxiety | C0860603 | Patient worries |
|  | Signs and symptoms of anxiety | C0860603 | Patient is fearful |
|  | Signs and symptoms of anxiety | C0860603 | Patient displays symptoms of anxiety |
|  | Signs and symptoms of anxiety | C0860603 | Patient is anxious |
|  | Signs and symptoms of anxiety | C0860603 | Currently experiences anxiety. |
|  | Generalized Anxiety Disorder | C0270549 | Diagnosed with Generalized Anxiety Disorder |
|  | Generalized Anxiety Disorder | C0270549 | Patient was diagnosed with Anxiety |
|  | family hx: Anxiety state | C0455385 | A family member had anxiety |
|  | family hx: Anxiety state | C0455385 | Patient's family member had anxiety |
|  | hx of anxiety state | C0455504 | Patient previously had anxiety |
|  | hx of anxiety state | C0455504 | Patient's had anxiety earlier |
| Depression | Major depressive disorder | C1269683 | Patient was diagnosed with depression |
|  | Major depressive disorder | C1269683 | Patient was diagnosed with MDD |
|  | Major depressive disorder | C1269683 | Patient was diagnosed with major depressive disorder |
|  | Major depressive disorder | C1269683 | Patient was diagnosed with dysthymia. |
|  | Family hx | C0455383 | Patient's family members had depression |
|  | Hx of Depression | C0455503 | Patient used to be depressed |
|  | Hx of Depression | C0455503 | Patient had depression earlier. |
|  | Symptoms of depression | C4716638 | Patient complains of low mood |
|  | Symptoms of depression | C4716638 | Patient felt depressed |
|  | Symptoms of depression | C4716638 | Patient appears depressed |
|  | Symptoms of depression | C4716638 | Patient displays symptoms of depression |
|  | Symptoms of depression | C4716638 | Positive for depression |
|  | Symptoms of depression | C4716638 | Patient is sad |
|  | Symptoms of depression | C4716638 | Patient feels down |
| Housing | Stably housed | C0020056 | The patient has proper housing |
|  | Stably housed | C0020056 | The patient is well housed |
|  | Stably housed | C0020056 | The patient has a home. |
|  | Homeless | C0237154 | The patient is homeless |
|  | Homeless | C0237154 | The patient has nowhere to live |
|  | Homeless | C0237154 | Patient is looking for housing |
|  | Homeless | C0237154 | Reports homelessness |
|  | Homeless | C0237154 | Homeless |
|  | Homeless | C0237154 | Patient doesn't have a home |
|  | Homeless | C0237154 | Patient lives on street |
|  | Homeless | C0237154 | Patient lives in car |
|  | Homeless | C0237154 | Patient lives in tent |
|  | Homeless | C0237154 | Patient is unsheltered |
|  | Homeless | C0237154 | Patient lives in encampment |
|  | Marginally housed | C5539303 | The housing is overcrowded |
|  | Marginally housed | C5539303 | Patient lives in an overcrowded place |
|  | Marginally housed | C5539303 | Patient has no home of their own |
|  | Marginally housed | C5539303 | Patient is marginally housed. |
|  | Subsidized housing | C5204673 | Patient lives in subsidized housing |
|  | Subsidized housing | C5204673 | Patient lives in public housing |
|  | Subsidized housing | C5204673 | Patient lives in sheltered housing |
|  | Housing unsuited to needs | C0578892 | The housing is unsuited for living |
|  | Housing unsuited to needs | C0578892 | Patient's housing presents health risks. |
|  | Lives in facility | C4510636 | The patient lives in a facility |
|  | NA | C0557202 | This describes patient's housing circumstances |
| Transportation | Has access to a car | C0425253 | Patient has access to a car |
|  | Has access to a car | C0425253 | Patient drives a car |
|  | Has access to a car | C0425253 | Patient drives a vehicle |
|  | Has access to public transport vehicle | C1446359 | Patient has access to public transportation |
|  | Has access to public transport vehicle | C1446359 | Patient rides public transportation |
|  | Transportation problems | C0425259 | Patient has issues getting around |
|  | Transportation problems | C0425259 | Patient experiences logistic problems |
|  | Transportation problems | C0425259 | Patient experiences transportation problems |
|  | NA | C3650743 | This describes patient's access to transportation |
| Finances | Financially secure | C0557160 | Patient is financially secure |
|  | Financially secure | C0557160 | Patient is in no financial distress |
|  | Financially secure | C0557160 | Patient is in no financial hardship |
|  | Financially secure | C0557160 | Patient is well off |
|  | Financially secure | C0557160 | Patient is wealthy. |
|  | Financial problem | C0549106 | Patient reports financial problem |
|  | Financial problem | C0549106 | Patient is financially distressed |
|  | Financial problem | C0549106 | Patient is afraid of a financial hardship |
|  | Financial problem | C0549106 | Patient is in financial hardship |
|  | Unable to afford medication | C4520569 | Patient is unable to afford medication |
|  | Unable to afford medication | C4520569 | The medication is too expensive |
|  | NA | C1287207 | This describes patient's financial situation |
|  | NA | C1287207 | This describes patient's employment |
| Social Support | NA | C1266850 | This describes patient's social circumstances. |
|  | At risk for loneliness | C1268628 | Patient is at risk for loneliness |
|  | Has social support | C3853122 | Patient has social support |
|  | Has social support | C3853122 | Patient keeps in touch with friends or relatives |
|  | Has social support | C3853122 | Patient is supported by family members |
|  | Has social support | C3853122 | Patient is a member of church |
|  | Has social support | C3853122 | Patient is a member of social club |
|  | Has social support | C3853122 | Patient interacts with relatives |
|  | Has social support | C3853122 | Patient interacts with children |
|  | Has social support | C3853122 | Patient is accompanied by a family member |
|  | Has social support | C3853122 | Patient has friends |
|  | Has social support | C3853122 | Relatives visit patient |
|  | Has social support | C3853122 | Patient came with a family member |
|  | Has social support | C3853122 | Patient is close with patient's family. |
|  | Social isolation | C0037421 | Patient is socially isolated |
|  | Social isolation | C0037421 | Doesn't have significant contact with or support from others. |
|  | Lives alone | C0439044 | Patient lives alone |
|  | Lives with | C0595998 | Patient lives with someone. |
|  | Personal relationship breakdown | C0524322 | Patient experienced a personal relationship breakdown |
|  | Personal relationship breakdown | C0524322 | Patient is no longer speaking to a family member |
|  | Personal relationship breakdown | C0524322 | Patient is no longer speaking to a friend. |
| Insurance status | Inadequate healthcare resources | C1828339 | Patient struggles to receive medical care |
|  | Inadequate healthcare resources | C1828339 | Patient has a limited insurance coverage |
|  | Inadequate healthcare resources | C1828339 | Patient's insurance denied some services |
|  | Inadequate healthcare resources | C1828339 | Patient lost insurance coverage. |
|  | NA | C1636784 | This describes patient's ability to access healthcare services or health insurance |
| Food | Able to obtain food | C0564281 | Patient is able to afford food |
|  | Provision of food | C4039510 | Patient is not able to afford food |
|  | Provision of food | C4039510 | Patient is insecure about getting enough food |
|  | Provision of food | C4039510 | Patient needs assistance with getting enough food |
|  | Provision of food | C4039510 | Patient receives food from a charity. |
|  | Fruit and vegetable intake | C1271941 | Patient eats fruits or vegetables |
|  | NA | C0577601 | There is information about patient's nutrition |
| Marital or partnership status | Separated | C4721417 | Patient is separated from partner |
|  | Separated | C4721417 | Patient broke up with the partner. |
|  | Divorced | C0086170 | Patient is divorced |
|  | Widowed | C1510465 | Patient is widowed |
|  | Partner | C0682323 | Patient has an intimate partner |
|  | Partner | C0682323 | Patient has a partner |
|  | Partner | C0682323 | Patient is seeing someone |
|  | Partner | C0682323 | Patient is in a relationship |
|  | Partner | C0682323 | Patient has a significant other. |
|  | Common law partnership | C0009448 | Patient is in common law partnership |
|  | Engaged to be married | C0425152 | Patient is engaged to be married |
|  | Married | C0555047 | Patient is married |
|  | Single person | C0037179 | Patient is single |
|  | Partner relationship problem | C0236858 | Patient experiences relationship problems |
|  | Partner relationship problem | C0236858 | Patient is a victim of domestic violence |
|  | NA | C0024819 | This describes patient's marital status |

Supplementary Table 3. Agreement (Cohen’s kappa) between individual annotators on document level

| Cohen’s kappa | M | R | S |
| --- | --- | --- | --- |
| E | 95.91% | 93.98% | 95.82% |
| M |  | 94.33% | 96.90% |
| R |  |  | 95.77% |

Supplementary Table 4. Agreement F_1_ between individual annotators on span level

| F_1_ | M | R | S |
| --- | --- | --- | --- |
| E | 93.41% | 88.83% | 91.08% |
| M |  | 90.26% | 95.03% |
| R |  |  | 88.60% |

Supplementary Table 5. Weighted average inter-rater agreement per label

| **label** | **span overlap F1, %** | **per-document Cohen's κ, %** |
| --- | --- | --- |
| Anxiety: GAD-7 | 84.21 | 99.74 |
| Anxiety: Generalized Anxiety Disorder | 74.51 | 97.52 |
| Anxiety: Signs and symptoms of anxiety | 89.41 | 96.94 |
| Anxiety: family hx: Anxiety state | 93.75 | 99.72 |
| Anxiety: hx of anxiety state | 93.31 | 98.23 |
| Anxiety: NA | 84.42 | 96.90 |
| Depression: Family hx: Depression | 98.34 | 99.43 |
| Depression: Major depressive disorder | 71.86 | 96.05 |
| Depression: PHQ-9 | 100.00 | 99.74 |
| Depression: Symptoms of depression | 85.36 | 95.52 |
| Depression: hx of Depression | 95.01 | 95.55 |
| Depression: NA | 35.29 | 97.00 |
| Finances: Financial problem | 100.00 | 99.62 |
| Finances: Financially secure | 100.00 | 99.66 |
| Finances: Unable to afford medication | 100.00 | 99.90 |
| Finances: NA | 100.00 | 97.73 |
| Food: Able to obtain food | 100.00 | 98.98 |
| Food: Fruit and vegetable intake | 100.00 | 100.00 |
| Food: Provision of food | 100.00 | 100.00 |
| Housing: Homeless | 100.00 | 99.82 |
| Housing: Housing unsuited to needs | 57.14 | 99.46 |
| Housing: Marginally housed | 36.36 | 99.66 |
| Housing: Stably housed | 97.83 | 90.74 |
| Housing: Subsidized housing | 100.00 | 100.00 |
| Housing: lives in facility | 85.71 | 99.52 |
| Housing: NA | 100.00 | 100.00 |
| Insurance Status: Inadequate healthcare resources | 98.63 | 99.94 |
| Insurance Status: NA | 98.67 | 99.12 |
| Marital_or_partnership_status: Divorced | 97.70 | 99.81 |
| Marital_or_partnership_status: Engaged to be married | 100.00 | 100.00 |
| Marital_or_partnership_status: Married | 98.05 | 98.06 |
| Marital_or_partnership_status: Partner | 96.03 | 98.27 |
| Marital_or_partnership_status: Partner relationship problem | 75.00 | 98.94 |
| Marital_or_partnership_status: Separated | 84.21 | 99.57 |
| Marital_or_partnership_status: Single person | 97.95 | 98.57 |
| Marital_or_partnership_status: Widowed | 98.32 | 99.85 |
| Marital_or_partnership_status: NA | 57.14 | 97.75 |
| Social Support : At risk for loneliness | 94.12 | 98.77 |
| Social Support : Has social support | 93.49 | 90.08 |
| Social Support : Lives alone | 98.77 | 98.85 |
| Social Support : Lives with | 97.25 | 97.43 |
| Social Support : Personal relationship breakdown | 100.00 | 98.46 |
| Social Support : Social isolation | 100.00 | 100.00 |
| Social Support : NA | 36.36 | 96.60 |
| Transportation: Has access to a car | 95.41 | 98.21 |
| Transportation: Has access to public transport vehicle | 93.75 | 99.04 |
| Transportation: Transportation problems | 100.00 | 99.83 |
| Transportation: NA | 78.26 | 98.66 |
| Pain Scores: Pain intensity rating scale, current | 89.08 | 93.88 |
| Pain Scores: Pain intensity rating scale, worst | 83.37 | 95.00 |
| Pain Scores: NA | 91.34 | 90.36 |

Supplementary Table 6. Detailed model performance metrics

| **label** | **level** | **entailment-RoBERTa** | **entailment-RoBERTa-mean** | **NER-hybrid** | **NER-CNN** | **NER-RoBERTa** | **cTAKES-SDoH** | **cTAKES-InfoCommons** | **cTAKES-default** |
| --- | --- | --- | --- | --- | --- | --- | --- | --- | --- |
| Anxiety | 1 | 67.08 | 38.58 | 91.71 | 90.82 | 90.64 | 63.98 | 68.84 | 74.46 |
| Depression | 1 | 83.77 | 36.52 | 91.95 | 90.26 | 90.48 | 68.58 | 66.90 | 70.39 |
| Marital_or partnership status | 1 | 88.82 | 39.32 | 89.78 | 92.38 | 92.33 | 63.80 | 41.61 | 45.15 |
| Pain_Scores | 1 | NA | NA | 94.93 | 93.80 | 95.82 | 12.17 | 0.00 | 0.00 |
| Housing | 1 | 60.43 | 10.49 | 61.80 | 64.47 | 67.66 | 7.01 | 6.93 | 0.00 |
| Social isolation | 1 | 76.93 | 22.07 | 47.41 | 61.35 | 68.80 | 28.06 | 29.25 | 22.25 |
| Transportation | 1 | 60.12 | 41.95 | 54.46 | 50.12 | 49.61 | 19.05 | 14.04 | 15.09 |
| Insurance status | 1 | 41.05 | 17.42 | 59.17 | 10.53 | 34.29 | 0.00 | 0.00 | 0.00 |
| Financial strain | 1 | 32.27 | 9.31 | 24.81 | 0.00 | 0.00 | 3.17 | 3.64 | 10.23 |
| Food | 1 | 5.41 | 5.28 | 0.00 | 0.00 | 0.00 | 0.00 | 0.00 | 0.00 |
| Anxiety | 2 | 43.39 | 47.44 | 73.51 | 67.62 | 72.57 | 33.19 | 30.60 | 27.21 |
| Depression | 2 | 42.71 | 27.23 | 62.27 | 58.42 | 67.96 | 29.07 | 15.58 | 8.67 |
| Marital_or_partnership_status | 2 | 77.18 | 65.50 | 83.46 | 87.17 | 85.42 | 72.94 | 44.07 | 37.60 |
| Pain_Scores | 2 | NA | NA | 84.86 | 84.51 | 87.77 | 10.36 | 0.00 | 0.00 |
| Housing | 2 | 62.73 | 47.74 | 52.62 | 55.01 | 56.92 | 0.68 | 0.60 | 0.00 |
| Social_isolation | 2 | 63.63 | 35.81 | 46.35 | 52.04 | 63.67 | 15.63 | 15.56 | 11.66 |
| Transportation | 2 | 62.23 | 74.97 | 48.23 | 41.41 | 43.41 | 20.23 | 13.22 | 14.43 |
| Insurance_status | 2 | 22.50 | 22.50 | 47.94 | 10.53 | 26.99 | 0.00 | 0.00 | 0.00 |
| Financial_strain | 2 | 48.68 | 19.93 | 0.00 | 0.00 | 0.00 | 1.74 | 1.67 | 6.28 |
| Food | 2 | 45.83 | 28.33 | 0.00 | 0.00 | 0.00 | NA | NA | NA |

Supplementary Table 7. Aggregated median metrics

| **method** | **f1-score: level-1** | **precision: level-1** | **recall: level-1** | **f1-score: level-2** | **precision: level-2** | **recall: level-2** |
| --- | --- | --- | --- | --- | --- | --- |
| entailment-RoBERTa | 76.04 | 67.44 | 89.53 | 58.15 | 54.96 | 73.09 |
| entailment-RoBERTa-mean | 30.65 | 63.32 | 20.46 | 42.35 | 56.73 | 40.12 |
| hybrid | 80.27 | 77.78 | 86.52 | 68.91 | 68.62 | 72.84 |
| CNN | 82.65 | 85.90 | 79.43 | 66.99 | 69.98 | 66.82 |
| cTAKES-SDoH | 47.11 | 42.87 | 57.20 | 34.32 | 44.10 | 35.24 |
| cTAKES-InfoCommons | 37.95 | 37.51 | 41.71 | 22.23 | 34.41 | 19.58 |
| cTAKES-default | 38.95 | 37.90 | 44.79 | 17.48 | 25.37 | 19.17 |

#### Supplementary Table 8A. Pair-wise comparison of model performance per first-level category (second-level metrics were preliminary aggregated within each cross-validation fold; paired two-sided t-test; positive difference indicates that the second model performed better, while negative indicates that the first model performed better). The “cTAKES” in this table refers to the SDOH configuration.

| **m1** | **m2** | **label** | **level** | **difference, % points** | **p.value** |
| --- | --- | --- | --- | --- | --- |
| entailment-RoBERTa-mean | entailment-RoBERTa | Anxiety | 1 | 30 | 3e-08 |
| entailment-RoBERTa-mean | entailment-RoBERTa | Anxiety | 2 | -5 | 0.2 |
| entailment-RoBERTa-mean | entailment-RoBERTa | Depression | 1 | 50 | 4e-10 |
| entailment-RoBERTa-mean | entailment-RoBERTa | Depression | 2 | 10 | 0.02 |
| entailment-RoBERTa-mean | entailment-RoBERTa | Marital_or_partnership_status | 1 | 50 | 2e-10 |
| entailment-RoBERTa-mean | entailment-RoBERTa | Marital_or_partnership_status | 2 | 10 | 9e-05 |
| entailment-RoBERTa-mean | entailment-RoBERTa | Housing | 1 | 50 | 2e-08 |
| entailment-RoBERTa-mean | entailment-RoBERTa | Housing | 2 | 20 | 1e-04 |
| entailment-RoBERTa-mean | entailment-RoBERTa | Social_isolation | 1 | 50 | 5e-07 |
| entailment-RoBERTa-mean | entailment-RoBERTa | Social_isolation | 2 | 30 | 5e-06 |
| entailment-RoBERTa-mean | entailment-RoBERTa | Transportation | 1 | 20 | 0.01 |
| entailment-RoBERTa-mean | entailment-RoBERTa | Transportation | 2 | -10 | 0.05 |
| entailment-RoBERTa-mean | entailment-RoBERTa | Insurance_status | 1 | 20 | 0.005 |
| entailment-RoBERTa-mean | entailment-RoBERTa | Insurance_status | 2 | 0 | 1 |
| entailment-RoBERTa-mean | entailment-RoBERTa | Financial_strain | 1 | 20 | 0.02 |
| entailment-RoBERTa-mean | entailment-RoBERTa | Financial_strain | 2 | 20 | 0.1 |
| entailment-RoBERTa-mean | entailment-RoBERTa | Food | 1 | 0 | 1 |
| entailment-RoBERTa-mean | entailment-RoBERTa | Food | 2 | 10 | 0.6 |
| NER-hybrid | entailment-RoBERTa | Anxiety | 1 | -20 | 3e-07 |
| NER-hybrid | entailment-RoBERTa | Anxiety | 2 | -30 | 7e-04 |
| NER-hybrid | entailment-RoBERTa | Depression | 1 | -8 | 0.002 |
| NER-hybrid | entailment-RoBERTa | Depression | 2 | -20 | 0.002 |
| NER-hybrid | entailment-RoBERTa | Marital_or_partnership_status | 1 | 0 | 1 |
| NER-hybrid | entailment-RoBERTa | Marital_or_partnership_status | 2 | -6 | 0.004 |
| NER-hybrid | entailment-RoBERTa | Housing | 1 | -2 | 0.5 |
| NER-hybrid | entailment-RoBERTa | Housing | 2 | 10 | 0.05 |
| NER-hybrid | entailment-RoBERTa | Social_isolation | 1 | 30 | 7e-06 |
| NER-hybrid | entailment-RoBERTa | Social_isolation | 2 | 20 | 2e-04 |
| NER-hybrid | entailment-RoBERTa | Transportation | 1 | 3 | 0.7 |
| NER-hybrid | entailment-RoBERTa | Transportation | 2 | 10 | 0.2 |
| NER-hybrid | entailment-RoBERTa | Insurance_status | 1 | -20 | 0.05 |
| NER-hybrid | entailment-RoBERTa | Insurance_status | 2 | -10 | 0.2 |
| NER-hybrid | entailment-RoBERTa | Financial_strain | 1 | 10 | 0.3 |
| NER-hybrid | entailment-RoBERTa | Financial_strain | 2 | 50 | 0.008 |
| NER-hybrid | entailment-RoBERTa | Food | 1 | 5 | 0.009 |
| NER-hybrid | entailment-RoBERTa | Food | 2 | 50 | 0.03 |
| NER-hybrid | entailment-RoBERTa-mean | Anxiety | 1 | -50 | 2e-11 |
| NER-hybrid | entailment-RoBERTa-mean | Anxiety | 2 | -20 | 0.003 |
| NER-hybrid | entailment-RoBERTa-mean | Depression | 1 | -60 | 4e-11 |
| NER-hybrid | entailment-RoBERTa-mean | Depression | 2 | -30 | 5e-05 |
| NER-hybrid | entailment-RoBERTa-mean | Marital_or_partnership_status | 1 | -50 | 1e-07 |
| NER-hybrid | entailment-RoBERTa-mean | Marital_or_partnership_status | 2 | -20 | 3e-06 |
| NER-hybrid | entailment-RoBERTa-mean | Housing | 1 | -50 | 6e-06 |
| NER-hybrid | entailment-RoBERTa-mean | Housing | 2 | -4 | 0.4 |
| NER-hybrid | entailment-RoBERTa-mean | Social_isolation | 1 | -30 | 2e-07 |
| NER-hybrid | entailment-RoBERTa-mean | Social_isolation | 2 | -9 | 0.004 |
| NER-hybrid | entailment-RoBERTa-mean | Transportation | 1 | -20 | 0.06 |
| NER-hybrid | entailment-RoBERTa-mean | Transportation | 2 | 20 | 0.03 |
| NER-hybrid | entailment-RoBERTa-mean | Insurance_status | 1 | -40 | 0.001 |
| NER-hybrid | entailment-RoBERTa-mean | Insurance_status | 2 | -10 | 0.2 |
| NER-hybrid | entailment-RoBERTa-mean | Financial_strain | 1 | -10 | 0.1 |
| NER-hybrid | entailment-RoBERTa-mean | Financial_strain | 2 | 20 | 0.03 |
| NER-hybrid | entailment-RoBERTa-mean | Food | 1 | 5 | 0.02 |
| NER-hybrid | entailment-RoBERTa-mean | Food | 2 | 40 | 0.04 |
| NER-CNN | entailment-RoBERTa | Anxiety | 1 | -20 | 3e-05 |
| NER-CNN | entailment-RoBERTa | Anxiety | 2 | -20 | 5e-05 |
| NER-CNN | entailment-RoBERTa | Depression | 1 | -8 | 0.002 |
| NER-CNN | entailment-RoBERTa | Depression | 2 | -20 | 0.002 |
| NER-CNN | entailment-RoBERTa | Marital_or_partnership_status | 1 | -4 | 0.01 |
| NER-CNN | entailment-RoBERTa | Marital_or_partnership_status | 2 | -9 | 7e-05 |
| NER-CNN | entailment-RoBERTa | Housing | 1 | -4 | 0.3 |
| NER-CNN | entailment-RoBERTa | Housing | 2 | 5 | 0.2 |
| NER-CNN | entailment-RoBERTa | Social_isolation | 1 | 20 | 0.003 |
| NER-CNN | entailment-RoBERTa | Social_isolation | 2 | 10 | 0.04 |
| NER-CNN | entailment-RoBERTa | Transportation | 1 | 20 | 0.1 |
| NER-CNN | entailment-RoBERTa | Transportation | 2 | 20 | 0.01 |
| NER-CNN | entailment-RoBERTa | Insurance_status | 1 | 20 | 0.2 |
| NER-CNN | entailment-RoBERTa | Insurance_status | 2 | 10 | 0.3 |
| NER-CNN | entailment-RoBERTa | Financial_strain | 1 | 30 | 0.005 |
| NER-CNN | entailment-RoBERTa | Financial_strain | 2 | 50 | 0.007 |
| NER-CNN | entailment-RoBERTa | Food | 1 | 6 | 0.02 |
| NER-CNN | entailment-RoBERTa | Food | 2 | 40 | 0.09 |
| NER-CNN | entailment-RoBERTa-mean | Anxiety | 1 | -50 | 1e-08 |
| NER-CNN | entailment-RoBERTa-mean | Anxiety | 2 | -20 | 0.001 |
| NER-CNN | entailment-RoBERTa-mean | Depression | 1 | -60 | 5e-11 |
| NER-CNN | entailment-RoBERTa-mean | Depression | 2 | -30 | 9e-06 |
| NER-CNN | entailment-RoBERTa-mean | Marital_or_partnership_status | 1 | -50 | 3e-08 |
| NER-CNN | entailment-RoBERTa-mean | Marital_or_partnership_status | 2 | -20 | 3e-07 |
| NER-CNN | entailment-RoBERTa-mean | Housing | 1 | -50 | 3e-06 |
| NER-CNN | entailment-RoBERTa-mean | Housing | 2 | -10 | 0.02 |
| NER-CNN | entailment-RoBERTa-mean | Social_isolation | 1 | -40 | 8e-05 |
| NER-CNN | entailment-RoBERTa-mean | Social_isolation | 2 | -20 | 0.008 |
| NER-CNN | entailment-RoBERTa-mean | Transportation | 1 | -3 | 0.7 |
| NER-CNN | entailment-RoBERTa-mean | Transportation | 2 | 40 | 0.002 |
| NER-CNN | entailment-RoBERTa-mean | Insurance_status | 1 | -6 | 0.6 |
| NER-CNN | entailment-RoBERTa-mean | Insurance_status | 2 | 10 | 0.3 |
| NER-CNN | entailment-RoBERTa-mean | Financial_strain | 1 | 10 | 0.02 |
| NER-CNN | entailment-RoBERTa-mean | Financial_strain | 2 | 20 | 0.03 |
| NER-CNN | entailment-RoBERTa-mean | Food | 1 | 6 | 0.02 |
| NER-CNN | entailment-RoBERTa-mean | Food | 2 | 40 | 0.08 |
| NER-CNN | NER-hybrid | Anxiety | 1 | 1 | 0.6 |
| NER-CNN | NER-hybrid | Anxiety | 2 | 3 | 0.5 |
| NER-CNN | NER-hybrid | Depression | 1 | 0 | 0.9 |
| NER-CNN | NER-hybrid | Depression | 2 | 2 | 0.7 |
| NER-CNN | NER-hybrid | Marital_or_partnership_status | 1 | -4 | 0.03 |
| NER-CNN | NER-hybrid | Marital_or_partnership_status | 2 | -3 | 0.1 |
| NER-CNN | NER-hybrid | Pain_Scores | 1 | 1 | 0.6 |
| NER-CNN | NER-hybrid | Pain_Scores | 2 | 4 | 0.6 |
| NER-CNN | NER-hybrid | Housing | 1 | -1 | 0.7 |
| NER-CNN | NER-hybrid | Housing | 2 | -6 | 0.3 |
| NER-CNN | NER-hybrid | Social_isolation | 1 | -10 | 0.03 |
| NER-CNN | NER-hybrid | Social_isolation | 2 | -7 | 0.2 |
| NER-CNN | NER-hybrid | Transportation | 1 | 10 | 0.3 |
| NER-CNN | NER-hybrid | Transportation | 2 | 20 | 0.2 |
| NER-CNN | NER-hybrid | Insurance_status | 1 | 40 | 0.02 |
| NER-CNN | NER-hybrid | Insurance_status | 2 | 20 | 0.06 |
| NER-CNN | NER-hybrid | Financial_strain | 1 | 20 | 0.01 |
| NER-CNN | NER-hybrid | Financial_strain | 2 | 1 | 0.4 |
| NER-CNN | NER-hybrid | Food | 1 | 1 | 0.4 |
| NER-CNN | NER-hybrid | Food | 2 | 0 | NaN |
| NER-RoBERTa | entailment-RoBERTa | Anxiety | 1 | -20 | 5e-07 |
| NER-RoBERTa | entailment-RoBERTa | Anxiety | 2 | -30 | 8e-05 |
| NER-RoBERTa | entailment-RoBERTa | Depression | 1 | -7 | 0.003 |
| NER-RoBERTa | entailment-RoBERTa | Depression | 2 | -30 | 3e-04 |
| NER-RoBERTa | entailment-RoBERTa | Marital_or_partnership_status | 1 | -4 | 9e-04 |
| NER-RoBERTa | entailment-RoBERTa | Marital_or_partnership_status | 2 | -8 | 2e-05 |
| NER-RoBERTa | entailment-RoBERTa | Housing | 1 | -4 | 0.4 |
| NER-RoBERTa | entailment-RoBERTa | Housing | 2 | 4 | 0.4 |
| NER-RoBERTa | entailment-RoBERTa | Social_isolation | 1 | 8 | 0.04 |
| NER-RoBERTa | entailment-RoBERTa | Social_isolation | 2 | 1 | 0.9 |
| NER-RoBERTa | entailment-RoBERTa | Transportation | 1 | 10 | 0.2 |
| NER-RoBERTa | entailment-RoBERTa | Transportation | 2 | 20 | 0.03 |
| NER-RoBERTa | entailment-RoBERTa | Insurance_status | 1 | 1 | 0.9 |
| NER-RoBERTa | entailment-RoBERTa | Insurance_status | 2 | 1 | 1 |
| NER-RoBERTa | entailment-RoBERTa | Financial_strain | 1 | 30 | 0.01 |
| NER-RoBERTa | entailment-RoBERTa | Financial_strain | 2 | 40 | 0.009 |
| NER-RoBERTa | entailment-RoBERTa | Food | 1 | 6 | 0.02 |
| NER-RoBERTa | entailment-RoBERTa | Food | 2 | 40 | 0.09 |
| NER-RoBERTa | entailment-RoBERTa-mean | Anxiety | 1 | -50 | 4e-11 |
| NER-RoBERTa | entailment-RoBERTa-mean | Anxiety | 2 | -20 | 6e-04 |
| NER-RoBERTa | entailment-RoBERTa-mean | Depression | 1 | -60 | 3e-10 |
| NER-RoBERTa | entailment-RoBERTa-mean | Depression | 2 | -40 | 2e-08 |
| NER-RoBERTa | entailment-RoBERTa-mean | Marital_or_partnership_status | 1 | -50 | 3e-10 |
| NER-RoBERTa | entailment-RoBERTa-mean | Marital_or_partnership_status | 2 | -20 | 8e-07 |
| NER-RoBERTa | entailment-RoBERTa-mean | Housing | 1 | -50 | 8e-05 |
| NER-RoBERTa | entailment-RoBERTa-mean | Housing | 2 | -10 | 0.06 |
| NER-RoBERTa | entailment-RoBERTa-mean | Social_isolation | 1 | -40 | 5e-06 |
| NER-RoBERTa | entailment-RoBERTa-mean | Social_isolation | 2 | -30 | 9e-05 |
| NER-RoBERTa | entailment-RoBERTa-mean | Transportation | 1 | -5 | 0.6 |
| NER-RoBERTa | entailment-RoBERTa-mean | Transportation | 2 | 30 | 0.007 |
| NER-RoBERTa | entailment-RoBERTa-mean | Insurance_status | 1 | -20 | 0.1 |
| NER-RoBERTa | entailment-RoBERTa-mean | Insurance_status | 2 | 1 | 1 |
| NER-RoBERTa | entailment-RoBERTa-mean | Financial_strain | 1 | 5 | 0.5 |
| NER-RoBERTa | entailment-RoBERTa-mean | Financial_strain | 2 | 20 | 0.06 |
| NER-RoBERTa | entailment-RoBERTa-mean | Food | 1 | 6 | 0.02 |
| NER-RoBERTa | entailment-RoBERTa-mean | Food | 2 | 40 | 0.08 |
| NER-RoBERTa | NER-hybrid | Anxiety | 1 | 0 | 0.8 |
| NER-RoBERTa | NER-hybrid | Anxiety | 2 | -2 | 0.7 |
| NER-RoBERTa | NER-hybrid | Depression | 1 | 1 | 0.7 |
| NER-RoBERTa | NER-hybrid | Depression | 2 | -6 | 0.2 |
| NER-RoBERTa | NER-hybrid | Marital_or_partnership_status | 1 | -4 | 0.03 |
| NER-RoBERTa | NER-hybrid | Marital_or_partnership_status | 2 | -2 | 0.3 |
| NER-RoBERTa | NER-hybrid | Pain_Scores | 1 | 0 | 0.9 |
| NER-RoBERTa | NER-hybrid | Pain_Scores | 2 | -2 | 0.6 |
| NER-RoBERTa | NER-hybrid | Housing | 1 | -2 | 0.7 |
| NER-RoBERTa | NER-hybrid | Housing | 2 | -7 | 0.3 |
| NER-RoBERTa | NER-hybrid | Social_isolation | 1 | -20 | 3e-04 |
| NER-RoBERTa | NER-hybrid | Social_isolation | 2 | -20 | 0.001 |
| NER-RoBERTa | NER-hybrid | Transportation | 1 | 10 | 0.3 |
| NER-RoBERTa | NER-hybrid | Transportation | 2 | 10 | 0.2 |
| NER-RoBERTa | NER-hybrid | Insurance_status | 1 | 20 | 0.1 |
| NER-RoBERTa | NER-hybrid | Insurance_status | 2 | 10 | 0.3 |
| NER-RoBERTa | NER-hybrid | Financial_strain | 1 | 20 | 0.08 |
| NER-RoBERTa | NER-hybrid | Financial_strain | 2 | -3 | 0.5 |
| NER-RoBERTa | NER-hybrid | Food | 1 | 1 | 0.4 |
| NER-RoBERTa | NER-hybrid | Food | 2 | 0 | NaN |
| NER-RoBERTa | NER-CNN | Anxiety | 1 | -1 | 0.5 |
| NER-RoBERTa | NER-CNN | Anxiety | 2 | -5 | 0.2 |
| NER-RoBERTa | NER-CNN | Depression | 1 | 0 | 0.8 |
| NER-RoBERTa | NER-CNN | Depression | 2 | -8 | 0.04 |
| NER-RoBERTa | NER-CNN | Marital_or_partnership_status | 1 | 0 | 0.8 |
| NER-RoBERTa | NER-CNN | Marital_or_partnership_status | 2 | 1 | 0.5 |
| NER-RoBERTa | NER-CNN | Pain_Scores | 1 | -1 | 0.5 |
| NER-RoBERTa | NER-CNN | Pain_Scores | 2 | -6 | 0.4 |
| NER-RoBERTa | NER-CNN | Housing | 1 | 0 | 0.9 |
| NER-RoBERTa | NER-CNN | Housing | 2 | -1 | 0.8 |
| NER-RoBERTa | NER-CNN | Social_isolation | 1 | -9 | 0.08 |
| NER-RoBERTa | NER-CNN | Social_isolation | 2 | -10 | 0.06 |
| NER-RoBERTa | NER-CNN | Transportation | 1 | -1 | 0.9 |
| NER-RoBERTa | NER-CNN | Transportation | 2 | -2 | 0.9 |
| NER-RoBERTa | NER-CNN | Insurance_status | 1 | -20 | 0.3 |
| NER-RoBERTa | NER-CNN | Insurance_status | 2 | -10 | 0.5 |
| NER-RoBERTa | NER-CNN | Financial_strain | 1 | -6 | 0.4 |
| NER-RoBERTa | NER-CNN | Financial_strain | 2 | -4 | 0.4 |
| NER-RoBERTa | NER-CNN | Food | 1 | 0 | NaN |
| NER-RoBERTa | NER-CNN | Food | 2 | 0 | NaN |
| cTAKES-SDoH | entailment-RoBERTa | Anxiety | 1 | 5 | 0.2 |
| cTAKES-SDoH | entailment-RoBERTa | Anxiety | 2 | 9 | 0.01 |
| cTAKES-SDoH | entailment-RoBERTa | Depression | 1 | 10 | 0.002 |
| cTAKES-SDoH | entailment-RoBERTa | Depression | 2 | 10 | 0.01 |
| cTAKES-SDoH | entailment-RoBERTa | Marital_or_partnership_status | 1 | 20 | 1e-04 |
| cTAKES-SDoH | entailment-RoBERTa | Marital_or_partnership_status | 2 | 4 | 0.03 |
| cTAKES-SDoH | entailment-RoBERTa | Housing | 1 | 50 | 4e-06 |
| cTAKES-SDoH | entailment-RoBERTa | Housing | 2 | 60 | 5e-09 |
| cTAKES-SDoH | entailment-RoBERTa | Social_isolation | 1 | 40 | 6e-06 |
| cTAKES-SDoH | entailment-RoBERTa | Social_isolation | 2 | 50 | 6e-08 |
| cTAKES-SDoH | entailment-RoBERTa | Transportation | 1 | 40 | 1e-04 |
| cTAKES-SDoH | entailment-RoBERTa | Transportation | 2 | 40 | 3e-05 |
| cTAKES-SDoH | entailment-RoBERTa | Insurance_status | 1 | 40 | 5e-04 |
| cTAKES-SDoH | entailment-RoBERTa | Insurance_status | 2 | 20 | 3e-04 |
| cTAKES-SDoH | entailment-RoBERTa | Financial_strain | 1 | 30 | 0.01 |
| cTAKES-SDoH | entailment-RoBERTa | Financial_strain | 2 | 40 | 0.01 |
| cTAKES-SDoH | entailment-RoBERTa | Food | 1 | 1 | 0.9 |
| cTAKES-SDoH | entailment-RoBERTa | Food | 2 | 40 | 0.1 |
| cTAKES-SDoH | entailment-RoBERTa-mean | Anxiety | 1 | -20 | 5e-04 |
| cTAKES-SDoH | entailment-RoBERTa-mean | Anxiety | 2 | 10 | 0.005 |
| cTAKES-SDoH | entailment-RoBERTa-mean | Depression | 1 | -40 | 1e-06 |
| cTAKES-SDoH | entailment-RoBERTa-mean | Depression | 2 | 1 | 0.7 |
| cTAKES-SDoH | entailment-RoBERTa-mean | Marital_or_partnership_status | 1 | -20 | 1e-04 |
| cTAKES-SDoH | entailment-RoBERTa-mean | Marital_or_partnership_status | 2 | -8 | 0.001 |
| cTAKES-SDoH | entailment-RoBERTa-mean | Housing | 1 | 0 | 1 |
| cTAKES-SDoH | entailment-RoBERTa-mean | Housing | 2 | 40 | 4e-06 |
| cTAKES-SDoH | entailment-RoBERTa-mean | Social_isolation | 1 | -8 | 0.08 |
| cTAKES-SDoH | entailment-RoBERTa-mean | Social_isolation | 2 | 20 | 2e-05 |
| cTAKES-SDoH | entailment-RoBERTa-mean | Transportation | 1 | 20 | 0.002 |
| cTAKES-SDoH | entailment-RoBERTa-mean | Transportation | 2 | 50 | 6e-06 |
| cTAKES-SDoH | entailment-RoBERTa-mean | Insurance_status | 1 | 10 | 0.001 |
| cTAKES-SDoH | entailment-RoBERTa-mean | Insurance_status | 2 | 20 | 3e-04 |
| cTAKES-SDoH | entailment-RoBERTa-mean | Financial_strain | 1 | 3 | 0.5 |
| cTAKES-SDoH | entailment-RoBERTa-mean | Financial_strain | 2 | 20 | 0.08 |
| cTAKES-SDoH | entailment-RoBERTa-mean | Food | 1 | 1 | 0.9 |
| cTAKES-SDoH | entailment-RoBERTa-mean | Food | 2 | 30 | 0.1 |
| cTAKES-SDoH | NER-hybrid | Anxiety | 1 | 30 | 1e-04 |
| cTAKES-SDoH | NER-hybrid | Anxiety | 2 | 40 | 2e-04 |
| cTAKES-SDoH | NER-hybrid | Depression | 1 | 20 | 1e-04 |
| cTAKES-SDoH | NER-hybrid | Depression | 2 | 30 | 6e-05 |
| cTAKES-SDoH | NER-hybrid | Marital_or_partnership_status | 1 | 20 | 3e-05 |
| cTAKES-SDoH | NER-hybrid | Marital_or_partnership_status | 2 | 10 | 3e-04 |
| cTAKES-SDoH | NER-hybrid | Pain_Scores | 1 | 80 | 7e-10 |
| cTAKES-SDoH | NER-hybrid | Pain_Scores | 2 | 70 | 7e-09 |
| cTAKES-SDoH | NER-hybrid | Housing | 1 | 50 | 8e-07 |
| cTAKES-SDoH | NER-hybrid | Housing | 2 | 50 | 1e-05 |
| cTAKES-SDoH | NER-hybrid | Social_isolation | 1 | 20 | 0.004 |
| cTAKES-SDoH | NER-hybrid | Social_isolation | 2 | 30 | 7e-07 |
| cTAKES-SDoH | NER-hybrid | Transportation | 1 | 40 | 7e-04 |
| cTAKES-SDoH | NER-hybrid | Transportation | 2 | 30 | 0.004 |
| cTAKES-SDoH | NER-hybrid | Insurance_status | 1 | 60 | 4e-04 |
| cTAKES-SDoH | NER-hybrid | Insurance_status | 2 | 40 | 0.004 |
| cTAKES-SDoH | NER-hybrid | Financial_strain | 1 | 20 | 0.07 |
| cTAKES-SDoH | NER-hybrid | Financial_strain | 2 | -5 | 0.2 |
| cTAKES-SDoH | NER-hybrid | Food | 1 | -5 | 0.5 |
| cTAKES-SDoH | NER-hybrid | Food | 2 | -5 | 0.4 |
| cTAKES-SDoH | NER-CNN | Anxiety | 1 | 30 | 7e-05 |
| cTAKES-SDoH | NER-CNN | Anxiety | 2 | 30 | 2e-06 |
| cTAKES-SDoH | NER-CNN | Depression | 1 | 20 | 1e-04 |
| cTAKES-SDoH | NER-CNN | Depression | 2 | 30 | 1e-05 |
| cTAKES-SDoH | NER-CNN | Marital_or_partnership_status | 1 | 30 | 1e-05 |
| cTAKES-SDoH | NER-CNN | Marital_or_partnership_status | 2 | 10 | 1e-05 |
| cTAKES-SDoH | NER-CNN | Pain_Scores | 1 | 80 | 8e-10 |
| cTAKES-SDoH | NER-CNN | Pain_Scores | 2 | 70 | 2e-05 |
| cTAKES-SDoH | NER-CNN | Housing | 1 | 50 | 6e-07 |
| cTAKES-SDoH | NER-CNN | Housing | 2 | 50 | 1e-07 |
| cTAKES-SDoH | NER-CNN | Social_isolation | 1 | 30 | 3e-04 |
| cTAKES-SDoH | NER-CNN | Social_isolation | 2 | 30 | 1e-04 |
| cTAKES-SDoH | NER-CNN | Transportation | 1 | 30 | 0.02 |
| cTAKES-SDoH | NER-CNN | Transportation | 2 | 20 | 0.06 |
| cTAKES-SDoH | NER-CNN | Insurance_status | 1 | 20 | 0.1 |
| cTAKES-SDoH | NER-CNN | Insurance_status | 2 | 20 | 0.1 |
| cTAKES-SDoH | NER-CNN | Financial_strain | 1 | -8 | 0.1 |
| cTAKES-SDoH | NER-CNN | Financial_strain | 2 | -7 | 0.1 |
| cTAKES-SDoH | NER-CNN | Food | 1 | -6 | 0.4 |
| cTAKES-SDoH | NER-CNN | Food | 2 | -5 | 0.4 |
| cTAKES-SDoH | NER-RoBERTa | Anxiety | 1 | 30 | 1e-04 |
| cTAKES-SDoH | NER-RoBERTa | Anxiety | 2 | 40 | 2e-05 |
| cTAKES-SDoH | NER-RoBERTa | Depression | 1 | 20 | 2e-04 |
| cTAKES-SDoH | NER-RoBERTa | Depression | 2 | 40 | 6e-09 |
| cTAKES-SDoH | NER-RoBERTa | Marital_or_partnership_status | 1 | 30 | 4e-05 |
| cTAKES-SDoH | NER-RoBERTa | Marital_or_partnership_status | 2 | 10 | 2e-05 |
| cTAKES-SDoH | NER-RoBERTa | Pain_Scores | 1 | 80 | 2e-10 |
| cTAKES-SDoH | NER-RoBERTa | Pain_Scores | 2 | 80 | 4e-08 |
| cTAKES-SDoH | NER-RoBERTa | Housing | 1 | 50 | 7e-06 |
| cTAKES-SDoH | NER-RoBERTa | Housing | 2 | 60 | 8e-06 |
| cTAKES-SDoH | NER-RoBERTa | Social_isolation | 1 | 40 | 2e-05 |
| cTAKES-SDoH | NER-RoBERTa | Social_isolation | 2 | 40 | 2e-06 |
| cTAKES-SDoH | NER-RoBERTa | Transportation | 1 | 30 | 0.02 |
| cTAKES-SDoH | NER-RoBERTa | Transportation | 2 | 20 | 0.07 |
| cTAKES-SDoH | NER-RoBERTa | Insurance_status | 1 | 40 | 0.02 |
| cTAKES-SDoH | NER-RoBERTa | Insurance_status | 2 | 20 | 0.06 |
| cTAKES-SDoH | NER-RoBERTa | Financial_strain | 1 | -2 | 0.8 |
| cTAKES-SDoH | NER-RoBERTa | Financial_strain | 2 | -2 | 0.7 |
| cTAKES-SDoH | NER-RoBERTa | Food | 1 | -6 | 0.4 |
| cTAKES-SDoH | NER-RoBERTa | Food | 2 | -5 | 0.4 |
| cTAKES-InfoCommons | entailment-RoBERTa | Anxiety | 1 | 0 | 1 |
| cTAKES-InfoCommons | entailment-RoBERTa | Anxiety | 2 | 10 | 0.003 |
| cTAKES-InfoCommons | entailment-RoBERTa | Depression | 1 | 20 | 1e-04 |
| cTAKES-InfoCommons | entailment-RoBERTa | Depression | 2 | 30 | 3e-04 |
| cTAKES-InfoCommons | entailment-RoBERTa | Marital_or_partnership_status | 1 | 50 | 1e-07 |
| cTAKES-InfoCommons | entailment-RoBERTa | Marital_or_partnership_status | 2 | 30 | 5e-08 |
| cTAKES-InfoCommons | entailment-RoBERTa | Housing | 1 | 50 | 4e-06 |
| cTAKES-InfoCommons | entailment-RoBERTa | Housing | 2 | 60 | 5e-09 |
| cTAKES-InfoCommons | entailment-RoBERTa | Social_isolation | 1 | 40 | 6e-06 |
| cTAKES-InfoCommons | entailment-RoBERTa | Social_isolation | 2 | 50 | 5e-08 |
| cTAKES-InfoCommons | entailment-RoBERTa | Transportation | 1 | 50 | 4e-05 |
| cTAKES-InfoCommons | entailment-RoBERTa | Transportation | 2 | 50 | 3e-06 |
| cTAKES-InfoCommons | entailment-RoBERTa | Insurance_status | 1 | 40 | 5e-04 |
| cTAKES-InfoCommons | entailment-RoBERTa | Insurance_status | 2 | 20 | 3e-04 |
| cTAKES-InfoCommons | entailment-RoBERTa | Financial_strain | 1 | 30 | 0.009 |
| cTAKES-InfoCommons | entailment-RoBERTa | Financial_strain | 2 | 40 | 0.01 |
| cTAKES-InfoCommons | entailment-RoBERTa | Food | 1 | 1 | 0.9 |
| cTAKES-InfoCommons | entailment-RoBERTa | Food | 2 | 40 | 0.1 |
| cTAKES-InfoCommons | entailment-RoBERTa-mean | Anxiety | 1 | -30 | 2e-04 |
| cTAKES-InfoCommons | entailment-RoBERTa-mean | Anxiety | 2 | 20 | 0.002 |
| cTAKES-InfoCommons | entailment-RoBERTa-mean | Depression | 1 | -30 | 2e-07 |
| cTAKES-InfoCommons | entailment-RoBERTa-mean | Depression | 2 | 10 | 2e-04 |
| cTAKES-InfoCommons | entailment-RoBERTa-mean | Marital_or_partnership_status | 1 | -2 | 0.3 |
| cTAKES-InfoCommons | entailment-RoBERTa-mean | Marital_or_partnership_status | 2 | 20 | 4e-07 |
| cTAKES-InfoCommons | entailment-RoBERTa-mean | Housing | 1 | 0 | 0.9 |
| cTAKES-InfoCommons | entailment-RoBERTa-mean | Housing | 2 | 40 | 4e-06 |
| cTAKES-InfoCommons | entailment-RoBERTa-mean | Social_isolation | 1 | -9 | 0.07 |
| cTAKES-InfoCommons | entailment-RoBERTa-mean | Social_isolation | 2 | 20 | 7e-05 |
| cTAKES-InfoCommons | entailment-RoBERTa-mean | Transportation | 1 | 30 | 2e-05 |
| cTAKES-InfoCommons | entailment-RoBERTa-mean | Transportation | 2 | 60 | 7e-07 |
| cTAKES-InfoCommons | entailment-RoBERTa-mean | Insurance_status | 1 | 10 | 0.001 |
| cTAKES-InfoCommons | entailment-RoBERTa-mean | Insurance_status | 2 | 20 | 3e-04 |
| cTAKES-InfoCommons | entailment-RoBERTa-mean | Financial_strain | 1 | 5 | 0.3 |
| cTAKES-InfoCommons | entailment-RoBERTa-mean | Financial_strain | 2 | 20 | 0.06 |
| cTAKES-InfoCommons | entailment-RoBERTa-mean | Food | 1 | 1 | 0.9 |
| cTAKES-InfoCommons | entailment-RoBERTa-mean | Food | 2 | 30 | 0.1 |
| cTAKES-InfoCommons | NER-hybrid | Anxiety | 1 | 20 | 3e-04 |
| cTAKES-InfoCommons | NER-hybrid | Anxiety | 2 | 40 | 8e-05 |
| cTAKES-InfoCommons | NER-hybrid | Depression | 1 | 20 | 5e-06 |
| cTAKES-InfoCommons | NER-hybrid | Depression | 2 | 50 | 6e-06 |
| cTAKES-InfoCommons | NER-hybrid | Marital_or_partnership_status | 1 | 50 | 1e-09 |
| cTAKES-InfoCommons | NER-hybrid | Marital_or_partnership_status | 2 | 40 | 2e-09 |
| cTAKES-InfoCommons | NER-hybrid | Pain_Scores | 1 | 100 | 4e-09 |
| cTAKES-InfoCommons | NER-hybrid | Pain_Scores | 2 | 80 | 1e-06 |
| cTAKES-InfoCommons | NER-hybrid | Housing | 1 | 50 | 8e-07 |
| cTAKES-InfoCommons | NER-hybrid | Housing | 2 | 50 | 1e-05 |
| cTAKES-InfoCommons | NER-hybrid | Social_isolation | 1 | 20 | 0.004 |
| cTAKES-InfoCommons | NER-hybrid | Social_isolation | 2 | 30 | 2e-06 |
| cTAKES-InfoCommons | NER-hybrid | Transportation | 1 | 40 | 4e-04 |
| cTAKES-InfoCommons | NER-hybrid | Transportation | 2 | 40 | 0.001 |
| cTAKES-InfoCommons | NER-hybrid | Insurance_status | 1 | 60 | 4e-04 |
| cTAKES-InfoCommons | NER-hybrid | Insurance_status | 2 | 40 | 0.004 |
| cTAKES-InfoCommons | NER-hybrid | Financial_strain | 1 | 20 | 0.04 |
| cTAKES-InfoCommons | NER-hybrid | Financial_strain | 2 | -4 | 0.3 |
| cTAKES-InfoCommons | NER-hybrid | Food | 1 | -5 | 0.5 |
| cTAKES-InfoCommons | NER-hybrid | Food | 2 | -5 | 0.4 |
| cTAKES-InfoCommons | NER-CNN | Anxiety | 1 | 20 | 3e-04 |
| cTAKES-InfoCommons | NER-CNN | Anxiety | 2 | 40 | 9e-07 |
| cTAKES-InfoCommons | NER-CNN | Depression | 1 | 20 | 8e-06 |
| cTAKES-InfoCommons | NER-CNN | Depression | 2 | 40 | 7e-07 |
| cTAKES-InfoCommons | NER-CNN | Marital_or_partnership_status | 1 | 50 | 2e-09 |
| cTAKES-InfoCommons | NER-CNN | Marital_or_partnership_status | 2 | 40 | 3e-10 |
| cTAKES-InfoCommons | NER-CNN | Pain_Scores | 1 | 90 | 4e-09 |
| cTAKES-InfoCommons | NER-CNN | Pain_Scores | 2 | 80 | 5e-05 |
| cTAKES-InfoCommons | NER-CNN | Housing | 1 | 50 | 6e-07 |
| cTAKES-InfoCommons | NER-CNN | Housing | 2 | 50 | 1e-07 |
| cTAKES-InfoCommons | NER-CNN | Social_isolation | 1 | 30 | 3e-04 |
| cTAKES-InfoCommons | NER-CNN | Social_isolation | 2 | 40 | 7e-05 |
| cTAKES-InfoCommons | NER-CNN | Transportation | 1 | 30 | 0.008 |
| cTAKES-InfoCommons | NER-CNN | Transportation | 2 | 20 | 0.01 |
| cTAKES-InfoCommons | NER-CNN | Insurance_status | 1 | 20 | 0.1 |
| cTAKES-InfoCommons | NER-CNN | Insurance_status | 2 | 20 | 0.1 |
| cTAKES-InfoCommons | NER-CNN | Financial_strain | 1 | -6 | 0.1 |
| cTAKES-InfoCommons | NER-CNN | Financial_strain | 2 | -5 | 0.1 |
| cTAKES-InfoCommons | NER-CNN | Food | 1 | -6 | 0.4 |
| cTAKES-InfoCommons | NER-CNN | Food | 2 | -5 | 0.4 |
| cTAKES-InfoCommons | NER-RoBERTa | Anxiety | 1 | 20 | 3e-04 |
| cTAKES-InfoCommons | NER-RoBERTa | Anxiety | 2 | 40 | 8e-06 |
| cTAKES-InfoCommons | NER-RoBERTa | Depression | 1 | 20 | 2e-05 |
| cTAKES-InfoCommons | NER-RoBERTa | Depression | 2 | 50 | 5e-10 |
| cTAKES-InfoCommons | NER-RoBERTa | Marital_or_partnership_status | 1 | 50 | 5e-08 |
| cTAKES-InfoCommons | NER-RoBERTa | Marital_or_partnership_status | 2 | 40 | 1e-09 |
| cTAKES-InfoCommons | NER-RoBERTa | Pain_Scores | 1 | 100 | 9e-09 |
| cTAKES-InfoCommons | NER-RoBERTa | Pain_Scores | 2 | 80 | 2e-06 |
| cTAKES-InfoCommons | NER-RoBERTa | Housing | 1 | 50 | 7e-06 |
| cTAKES-InfoCommons | NER-RoBERTa | Housing | 2 | 60 | 8e-06 |
| cTAKES-InfoCommons | NER-RoBERTa | Social_isolation | 1 | 40 | 2e-05 |
| cTAKES-InfoCommons | NER-RoBERTa | Social_isolation | 2 | 50 | 1e-06 |
| cTAKES-InfoCommons | NER-RoBERTa | Transportation | 1 | 30 | 0.007 |
| cTAKES-InfoCommons | NER-RoBERTa | Transportation | 2 | 30 | 0.02 |
| cTAKES-InfoCommons | NER-RoBERTa | Insurance_status | 1 | 40 | 0.02 |
| cTAKES-InfoCommons | NER-RoBERTa | Insurance_status | 2 | 20 | 0.06 |
| cTAKES-InfoCommons | NER-RoBERTa | Financial_strain | 1 | 0 | 1 |
| cTAKES-InfoCommons | NER-RoBERTa | Financial_strain | 2 | 0 | 0.9 |
| cTAKES-InfoCommons | NER-RoBERTa | Food | 1 | -6 | 0.4 |
| cTAKES-InfoCommons | NER-RoBERTa | Food | 2 | -5 | 0.4 |
| cTAKES-InfoCommons | cTAKES-SDoH | Anxiety | 1 | -5 | 0.3 |
| cTAKES-InfoCommons | cTAKES-SDoH | Anxiety | 2 | 3 | 0.4 |
| cTAKES-InfoCommons | cTAKES-SDoH | Depression | 1 | 2 | 0.4 |
| cTAKES-InfoCommons | cTAKES-SDoH | Depression | 2 | 10 | 1e-04 |
| cTAKES-InfoCommons | cTAKES-SDoH | Marital_or_partnership_status | 1 | 20 | 3e-05 |
| cTAKES-InfoCommons | cTAKES-SDoH | Marital_or_partnership_status | 2 | 30 | 1e-08 |
| cTAKES-InfoCommons | cTAKES-SDoH | Pain_Scores | 1 | 10 | 0.002 |
| cTAKES-InfoCommons | cTAKES-SDoH | Pain_Scores | 2 | 10 | 0.008 |
| cTAKES-InfoCommons | cTAKES-SDoH | Housing | 1 | 0 | 1 |
| cTAKES-InfoCommons | cTAKES-SDoH | Housing | 2 | 0 | 1 |
| cTAKES-InfoCommons | cTAKES-SDoH | Social_isolation | 1 | 0 | 1 |
| cTAKES-InfoCommons | cTAKES-SDoH | Social_isolation | 2 | 0 | 0.9 |
| cTAKES-InfoCommons | cTAKES-SDoH | Transportation | 1 | 6 | 0.3 |
| cTAKES-InfoCommons | cTAKES-SDoH | Transportation | 2 | 7 | 0.3 |
| cTAKES-InfoCommons | cTAKES-SDoH | Insurance_status | 1 | 0 | 1 |
| cTAKES-InfoCommons | cTAKES-SDoH | Insurance_status | 2 | 0 | 1 |
| cTAKES-InfoCommons | cTAKES-SDoH | Financial_strain | 1 | 2 | 0.8 |
| cTAKES-InfoCommons | cTAKES-SDoH | Financial_strain | 2 | 2 | 0.7 |
| cTAKES-InfoCommons | cTAKES-SDoH | Food | 1 | 0 | 1 |
| cTAKES-InfoCommons | cTAKES-SDoH | Food | 2 | 0 | 1 |
| cTAKES-default | entailment-RoBERTa | Anxiety | 1 | -5 | 0.2 |
| cTAKES-default | entailment-RoBERTa | Anxiety | 2 | 20 | 2e-04 |
| cTAKES-default | entailment-RoBERTa | Depression | 1 | 10 | 0.001 |
| cTAKES-default | entailment-RoBERTa | Depression | 2 | 30 | 7e-05 |
| cTAKES-default | entailment-RoBERTa | Marital_or_partnership_status | 1 | 40 | 1e-07 |
| cTAKES-default | entailment-RoBERTa | Marital_or_partnership_status | 2 | 40 | 1e-08 |
| cTAKES-default | entailment-RoBERTa | Housing | 1 | 60 | 6e-06 |
| cTAKES-default | entailment-RoBERTa | Housing | 2 | 60 | 7e-09 |
| cTAKES-default | entailment-RoBERTa | Social_isolation | 1 | 60 | 2e-08 |
| cTAKES-default | entailment-RoBERTa | Social_isolation | 2 | 50 | 4e-08 |
| cTAKES-default | entailment-RoBERTa | Transportation | 1 | 40 | 8e-05 |
| cTAKES-default | entailment-RoBERTa | Transportation | 2 | 50 | 2e-06 |
| cTAKES-default | entailment-RoBERTa | Insurance_status | 1 | 40 | 7e-04 |
| cTAKES-default | entailment-RoBERTa | Insurance_status | 2 | 30 | 6e-04 |
| cTAKES-default | entailment-RoBERTa | Financial_strain | 1 | 30 | 0.01 |
| cTAKES-default | entailment-RoBERTa | Financial_strain | 2 | 40 | 0.01 |
| cTAKES-default | entailment-RoBERTa | Food | 1 | 6 | 0.02 |
| cTAKES-default | entailment-RoBERTa | Food | 2 | 40 | 0.09 |
| cTAKES-default | entailment-RoBERTa-mean | Anxiety | 1 | -30 | 1e-04 |
| cTAKES-default | entailment-RoBERTa-mean | Anxiety | 2 | 20 | 4e-04 |
| cTAKES-default | entailment-RoBERTa-mean | Depression | 1 | -30 | 1e-06 |
| cTAKES-default | entailment-RoBERTa-mean | Depression | 2 | 20 | 8e-06 |
| cTAKES-default | entailment-RoBERTa-mean | Marital_or_partnership_status | 1 | -5 | 0.02 |
| cTAKES-default | entailment-RoBERTa-mean | Marital_or_partnership_status | 2 | 30 | 2e-08 |
| cTAKES-default | entailment-RoBERTa-mean | Housing | 1 | 6 | 0.2 |
| cTAKES-default | entailment-RoBERTa-mean | Housing | 2 | 40 | 4e-06 |
| cTAKES-default | entailment-RoBERTa-mean | Social_isolation | 1 | 1 | 0.6 |
| cTAKES-default | entailment-RoBERTa-mean | Social_isolation | 2 | 20 | 2e-06 |
| cTAKES-default | entailment-RoBERTa-mean | Transportation | 1 | 30 | 3e-05 |
| cTAKES-default | entailment-RoBERTa-mean | Transportation | 2 | 60 | 1e-07 |
| cTAKES-default | entailment-RoBERTa-mean | Insurance_status | 1 | 20 | 0.002 |
| cTAKES-default | entailment-RoBERTa-mean | Insurance_status | 2 | 30 | 6e-04 |
| cTAKES-default | entailment-RoBERTa-mean | Financial_strain | 1 | 3 | 0.5 |
| cTAKES-default | entailment-RoBERTa-mean | Financial_strain | 2 | 20 | 0.09 |
| cTAKES-default | entailment-RoBERTa-mean | Food | 1 | 6 | 0.02 |
| cTAKES-default | entailment-RoBERTa-mean | Food | 2 | 40 | 0.08 |
| cTAKES-default | NER-hybrid | Anxiety | 1 | 20 | 0.002 |
| cTAKES-default | NER-hybrid | Anxiety | 2 | 40 | 4e-05 |
| cTAKES-default | NER-hybrid | Depression | 1 | 20 | 6e-05 |
| cTAKES-default | NER-hybrid | Depression | 2 | 50 | 3e-06 |
| cTAKES-default | NER-hybrid | Marital_or_partnership_status | 1 | 40 | 2e-09 |
| cTAKES-default | NER-hybrid | Marital_or_partnership_status | 2 | 50 | 3e-10 |
| cTAKES-default | NER-hybrid | Pain_Scores | 1 | 100 | 4e-09 |
| cTAKES-default | NER-hybrid | Pain_Scores | 2 | 80 | 1e-06 |
| cTAKES-default | NER-hybrid | Housing | 1 | 60 | 8e-07 |
| cTAKES-default | NER-hybrid | Housing | 2 | 50 | 1e-05 |
| cTAKES-default | NER-hybrid | Social_isolation | 1 | 30 | 1e-05 |
| cTAKES-default | NER-hybrid | Social_isolation | 2 | 30 | 1e-07 |
| cTAKES-default | NER-hybrid | Transportation | 1 | 40 | 6e-04 |
| cTAKES-default | NER-hybrid | Transportation | 2 | 40 | 0.002 |
| cTAKES-default | NER-hybrid | Insurance_status | 1 | 60 | 5e-04 |
| cTAKES-default | NER-hybrid | Insurance_status | 2 | 40 | 0.004 |
| cTAKES-default | NER-hybrid | Financial_strain | 1 | 10 | 0.07 |
| cTAKES-default | NER-hybrid | Financial_strain | 2 | -6 | 0.1 |
| cTAKES-default | NER-hybrid | Food | 1 | 1 | 0.4 |
| cTAKES-default | NER-hybrid | Food | 2 | 0 | NaN |
| cTAKES-default | NER-CNN | Anxiety | 1 | 20 | 0.003 |
| cTAKES-default | NER-CNN | Anxiety | 2 | 40 | 3e-07 |
| cTAKES-default | NER-CNN | Depression | 1 | 20 | 8e-05 |
| cTAKES-default | NER-CNN | Depression | 2 | 50 | 3e-07 |
| cTAKES-default | NER-CNN | Marital_or_partnership_status | 1 | 50 | 2e-09 |
| cTAKES-default | NER-CNN | Marital_or_partnership_status | 2 | 50 | 7e-11 |
| cTAKES-default | NER-CNN | Pain_Scores | 1 | 90 | 4e-09 |
| cTAKES-default | NER-CNN | Pain_Scores | 2 | 80 | 5e-05 |
| cTAKES-default | NER-CNN | Housing | 1 | 60 | 7e-07 |
| cTAKES-default | NER-CNN | Housing | 2 | 60 | 1e-07 |
| cTAKES-default | NER-CNN | Social_isolation | 1 | 40 | 8e-06 |
| cTAKES-default | NER-CNN | Social_isolation | 2 | 40 | 6e-05 |
| cTAKES-default | NER-CNN | Transportation | 1 | 30 | 0.01 |
| cTAKES-default | NER-CNN | Transportation | 2 | 20 | 0.02 |
| cTAKES-default | NER-CNN | Insurance_status | 1 | 20 | 0.1 |
| cTAKES-default | NER-CNN | Insurance_status | 2 | 20 | 0.1 |
| cTAKES-default | NER-CNN | Financial_strain | 1 | -8 | 0.03 |
| cTAKES-default | NER-CNN | Financial_strain | 2 | -7 | 0.06 |
| cTAKES-default | NER-CNN | Food | 1 | 0 | NaN |
| cTAKES-default | NER-CNN | Food | 2 | 0 | NaN |
| cTAKES-default | NER-RoBERTa | Anxiety | 1 | 20 | 0.002 |
| cTAKES-default | NER-RoBERTa | Anxiety | 2 | 40 | 4e-06 |
| cTAKES-default | NER-RoBERTa | Depression | 1 | 20 | 1e-04 |
| cTAKES-default | NER-RoBERTa | Depression | 2 | 60 | 1e-10 |
| cTAKES-default | NER-RoBERTa | Marital_or_partnership_status | 1 | 50 | 5e-08 |
| cTAKES-default | NER-RoBERTa | Marital_or_partnership_status | 2 | 50 | 2e-10 |
| cTAKES-default | NER-RoBERTa | Pain_Scores | 1 | 100 | 9e-09 |
| cTAKES-default | NER-RoBERTa | Pain_Scores | 2 | 80 | 2e-06 |
| cTAKES-default | NER-RoBERTa | Housing | 1 | 60 | 3e-06 |
| cTAKES-default | NER-RoBERTa | Housing | 2 | 60 | 7e-06 |
| cTAKES-default | NER-RoBERTa | Social_isolation | 1 | 50 | 2e-07 |
| cTAKES-default | NER-RoBERTa | Social_isolation | 2 | 50 | 2e-06 |
| cTAKES-default | NER-RoBERTa | Transportation | 1 | 30 | 0.009 |
| cTAKES-default | NER-RoBERTa | Transportation | 2 | 20 | 0.03 |
| cTAKES-default | NER-RoBERTa | Insurance_status | 1 | 40 | 0.02 |
| cTAKES-default | NER-RoBERTa | Insurance_status | 2 | 30 | 0.05 |
| cTAKES-default | NER-RoBERTa | Financial_strain | 1 | -2 | 0.8 |
| cTAKES-default | NER-RoBERTa | Financial_strain | 2 | -3 | 0.6 |
| cTAKES-default | NER-RoBERTa | Food | 1 | 0 | NaN |
| cTAKES-default | NER-RoBERTa | Food | 2 | 0 | NaN |
| cTAKES-default | cTAKES-SDoH | Anxiety | 1 | -10 | 0.06 |
| cTAKES-default | cTAKES-SDoH | Anxiety | 2 | 8 | 0.01 |
| cTAKES-default | cTAKES-SDoH | Depression | 1 | 1 | 0.8 |
| cTAKES-default | cTAKES-SDoH | Depression | 2 | 20 | 4e-06 |
| cTAKES-default | cTAKES-SDoH | Marital_or_partnership_status | 1 | 20 | 1e-04 |
| cTAKES-default | cTAKES-SDoH | Marital_or_partnership_status | 2 | 40 | 1e-09 |
| cTAKES-default | cTAKES-SDoH | Pain_Scores | 1 | 10 | 0.002 |
| cTAKES-default | cTAKES-SDoH | Pain_Scores | 2 | 10 | 0.008 |
| cTAKES-default | cTAKES-SDoH | Housing | 1 | 6 | 0.3 |
| cTAKES-default | cTAKES-SDoH | Housing | 2 | 1 | 0.8 |
| cTAKES-default | cTAKES-SDoH | Social_isolation | 1 | 10 | 0.06 |
| cTAKES-default | cTAKES-SDoH | Social_isolation | 2 | 5 | 0.06 |
| cTAKES-default | cTAKES-SDoH | Transportation | 1 | 4 | 0.4 |
| cTAKES-default | cTAKES-SDoH | Transportation | 2 | 5 | 0.4 |
| cTAKES-default | cTAKES-SDoH | Insurance_status | 1 | 1 | 0.4 |
| cTAKES-default | cTAKES-SDoH | Insurance_status | 2 | 1 | 0.4 |
| cTAKES-default | cTAKES-SDoH | Financial_strain | 1 | -1 | 0.9 |
| cTAKES-default | cTAKES-SDoH | Financial_strain | 2 | 0 | 0.9 |
| cTAKES-default | cTAKES-SDoH | Food | 1 | 6 | 0.4 |
| cTAKES-default | cTAKES-SDoH | Food | 2 | 5 | 0.4 |
| cTAKES-default | cTAKES-InfoCommons | Anxiety | 1 | -5 | 0.3 |
| cTAKES-default | cTAKES-InfoCommons | Anxiety | 2 | 5 | 0.08 |
| cTAKES-default | cTAKES-InfoCommons | Depression | 1 | -1 | 0.6 |
| cTAKES-default | cTAKES-InfoCommons | Depression | 2 | 6 | 0.02 |
| cTAKES-default | cTAKES-InfoCommons | Marital_or_partnership_status | 1 | -3 | 0.2 |
| cTAKES-default | cTAKES-InfoCommons | Marital_or_partnership_status | 2 | 8 | 0.002 |
| cTAKES-default | cTAKES-InfoCommons | Pain_Scores | 1 | 0 | 0.4 |
| cTAKES-default | cTAKES-InfoCommons | Pain_Scores | 2 | 0 | NaN |
| cTAKES-default | cTAKES-InfoCommons | Housing | 1 | 6 | 0.3 |
| cTAKES-default | cTAKES-InfoCommons | Housing | 2 | 1 | 0.8 |
| cTAKES-default | cTAKES-InfoCommons | Social_isolation | 1 | 10 | 0.06 |
| cTAKES-default | cTAKES-InfoCommons | Social_isolation | 2 | 4 | 0.1 |
| cTAKES-default | cTAKES-InfoCommons | Transportation | 1 | -2 | 0.6 |
| cTAKES-default | cTAKES-InfoCommons | Transportation | 2 | -2 | 0.8 |
| cTAKES-default | cTAKES-InfoCommons | Insurance_status | 1 | 1 | 0.4 |
| cTAKES-default | cTAKES-InfoCommons | Insurance_status | 2 | 1 | 0.4 |
| cTAKES-default | cTAKES-InfoCommons | Financial_strain | 1 | -2 | 0.6 |
| cTAKES-default | cTAKES-InfoCommons | Financial_strain | 2 | -2 | 0.6 |
| cTAKES-default | cTAKES-InfoCommons | Food | 1 | 6 | 0.4 |
| cTAKES-default | cTAKES-InfoCommons | Food | 2 | 5 | 0.4 |

Supplementary Table 8B. Detailed model comparison across labels at the second level

| **model 1** | **model 2** | **label** | **difference, % points** | **p-value** |
| --- | --- | --- | --- | --- |
| cTAKES-SDoH | CNN | Anxiety: family hx: Anxiety state | -21 | 0.4 |
| cTAKES-SDoH | CNN | Anxiety: Generalized Anxiety Disorder | 16 | 0.06 |
| cTAKES-SDoH | CNN | Anxiety: hx of anxiety state | 27 | 0.005 |
| cTAKES-SDoH | CNN | Anxiety: NA | 74 | 9e-06 |
| cTAKES-SDoH | CNN | Anxiety: Signs and symptoms of anxiety | 58 | 3e-07 |
| cTAKES-SDoH | CNN | Depression: Family hx: Depression | 16 | 0.06 |
| cTAKES-SDoH | CNN | Depression: hx of Depression | 43 | 8e-06 |
| cTAKES-SDoH | CNN | Depression: Major depressive disorder | 4 | 0.4 |
| cTAKES-SDoH | CNN | Depression: Symptoms of depression | 52 | 2e-07 |
| cTAKES-SDoH | CNN | Housing: Homeless | -22 | 0.6 |
| cTAKES-SDoH | CNN | Insurance_status: NA | 17 | 0.1 |
| cTAKES-SDoH | CNN | Marital_or_partnership_status: Divorced | 10 | 0.05 |
| cTAKES-SDoH | CNN | Marital_or_partnership_status: Married | 23 | 5e-05 |
| cTAKES-SDoH | CNN | Marital_or_partnership_status: Partner | 40 | 0.02 |
| cTAKES-SDoH | CNN | Marital_or_partnership_status: Single person | 29 | 0.002 |
| cTAKES-SDoH | CNN | Marital_or_partnership_status: Widowed | 46 | 0.004 |
| cTAKES-SDoH | CNN | pain_and_disability: NA | 80 | 1e-07 |
| cTAKES-SDoH | CNN | Social_support: Has social support | 29 | 0.002 |
| cTAKES-SDoH | CNN | Social_support: Lives alone | 18 | 0.06 |
| cTAKES-SDoH | CNN | Social_support: Lives with | 59 | 7e-07 |
| cTAKES-SDoH | CNN | Transportation: Has access to a car | 10 | 0.1 |
| cTAKES-SDoH | CNN | Transportation: Transportation problems | 67 | 0.006 |
| cTAKES-SDoH | hybrid | Anxiety: family hx: Anxiety state | 12 | 0.7 |
| cTAKES-SDoH | hybrid | Anxiety: Generalized Anxiety Disorder | 32 | 0.02 |
| cTAKES-SDoH | hybrid | Anxiety: hx of anxiety state | 29 | 0.003 |
| cTAKES-SDoH | hybrid | Anxiety: NA | 79 | 2e-05 |
| cTAKES-SDoH | hybrid | Anxiety: Signs and symptoms of anxiety | 59 | 1e-05 |
| cTAKES-SDoH | hybrid | Depression: Family hx: Depression | 23 | 0.01 |
| cTAKES-SDoH | hybrid | Depression: hx of Depression | 44 | 5e-06 |
| cTAKES-SDoH | hybrid | Depression: Major depressive disorder | 20 | 0.07 |
| cTAKES-SDoH | hybrid | Depression: NA | 8 | 0.2 |
| cTAKES-SDoH | hybrid | Depression: Symptoms of depression | 54 | 6e-07 |
| cTAKES-SDoH | hybrid | Financial_strain: Financial problem | 8 | 0.6 |
| cTAKES-SDoH | hybrid | Financial_strain: NA | 11 | 0.4 |
| cTAKES-SDoH | hybrid | Housing: Homeless | -23 | 0.4 |
| cTAKES-SDoH | hybrid | Insurance_status: NA | 45 | 0.009 |
| cTAKES-SDoH | hybrid | Marital_or_partnership_status: Divorced | 4 | 0.4 |
| cTAKES-SDoH | hybrid | Marital_or_partnership_status: Married | 20 | 1e-04 |
| cTAKES-SDoH | hybrid | Marital_or_partnership_status: NA | 21 | 0.1 |
| cTAKES-SDoH | hybrid | Marital_or_partnership_status: Partner | 43 | 0.02 |
| cTAKES-SDoH | hybrid | Marital_or_partnership_status: Partner relationship problem | 12 | 0.4 |
| cTAKES-SDoH | hybrid | Marital_or_partnership_status: Single person | 24 | 0.003 |
| cTAKES-SDoH | hybrid | Marital_or_partnership_status: Widowed | -7 | 0.6 |
| cTAKES-SDoH | hybrid | pain_and_disability: NA | 82 | 1e-09 |
| cTAKES-SDoH | hybrid | Social_support: At risk for loneliness | 11 | 0.3 |
| cTAKES-SDoH | hybrid | Social_support: Has social support | 31 | 1e-06 |
| cTAKES-SDoH | hybrid | Social_support: Lives alone | 10 | 0.3 |
| cTAKES-SDoH | hybrid | Social_support: Lives with | 42 | 9e-07 |
| cTAKES-SDoH | hybrid | Transportation: Has access to a car | 29 | 0.009 |
| cTAKES-SDoH | hybrid | Transportation: Has access to public transport vehicle | 30 | 0.02 |
| cTAKES-SDoH | hybrid | Transportation: Transportation problems | 69 | 0.005 |
| cTAKES-SDoH | entailment-RoBERTa-mean | Anxiety: family hx: Anxiety state | 25 | 0.4 |
| cTAKES-SDoH | entailment-RoBERTa-mean | Anxiety: Generalized Anxiety Disorder | 7 | 0.2 |
| cTAKES-SDoH | entailment-RoBERTa-mean | Anxiety: hx of anxiety state | -1 | 0.9 |
| cTAKES-SDoH | entailment-RoBERTa-mean | Anxiety: Signs and symptoms of anxiety | 38 | 4e-07 |
| cTAKES-SDoH | entailment-RoBERTa-mean | Depression: Family hx: Depression | -18 | 0.2 |
| cTAKES-SDoH | entailment-RoBERTa-mean | Depression: hx of Depression | -10 | 0.3 |
| cTAKES-SDoH | entailment-RoBERTa-mean | Depression: Major depressive disorder | 3 | 0.2 |
| cTAKES-SDoH | entailment-RoBERTa-mean | Depression: Symptoms of depression | 25 | 1e-04 |
| cTAKES-SDoH | entailment-RoBERTa-mean | Financial_strain: Financial problem | 34 | 0.04 |
| cTAKES-SDoH | entailment-RoBERTa-mean | Financial_strain: NA | 21 | 0.1 |
| cTAKES-SDoH | entailment-RoBERTa-mean | Housing: Homeless | 1 | 1 |
| cTAKES-SDoH | entailment-RoBERTa-mean | Insurance_status: NA | 38 | 0.03 |
| cTAKES-SDoH | entailment-RoBERTa-mean | Marital_or_partnership_status: Divorced | -23 | 0.009 |
| cTAKES-SDoH | entailment-RoBERTa-mean | Marital_or_partnership_status: Married | 23 | 1e-04 |
| cTAKES-SDoH | entailment-RoBERTa-mean | Marital_or_partnership_status: NA | 43 | 0.05 |
| cTAKES-SDoH | entailment-RoBERTa-mean | Marital_or_partnership_status: Partner | 1 | 1 |
| cTAKES-SDoH | entailment-RoBERTa-mean | Marital_or_partnership_status: Partner relationship problem | 14 | 0.3 |
| cTAKES-SDoH | entailment-RoBERTa-mean | Marital_or_partnership_status: Single person | 19 | 0.01 |
| cTAKES-SDoH | entailment-RoBERTa-mean | Marital_or_partnership_status: Widowed | 33 | 0.02 |
| cTAKES-SDoH | entailment-RoBERTa-mean | Social_support: Has social support | 26 | 2e-06 |
| cTAKES-SDoH | entailment-RoBERTa-mean | Social_support: Lives alone | 31 | 0.005 |
| cTAKES-SDoH | entailment-RoBERTa-mean | Social_support: Lives with | 69 | 9e-09 |
| cTAKES-SDoH | entailment-RoBERTa-mean | Social_support: NA | 21 | 0.2 |
| cTAKES-SDoH | entailment-RoBERTa-mean | Social_support: Personal relationship breakdown | 4 | 0.4 |
| cTAKES-SDoH | entailment-RoBERTa-mean | Transportation: Has access to a car | 63 | 1e-06 |
| cTAKES-SDoH | entailment-RoBERTa-mean | Transportation: Has access to public transport vehicle | 61 | 5e-05 |
| cTAKES-SDoH | entailment-RoBERTa-mean | Transportation: Transportation problems | 44 | 0.002 |
| cTAKES-SDoH | entailment-RoBERTa | Anxiety: family hx: Anxiety state | -8 | 0.8 |
| cTAKES-SDoH | entailment-RoBERTa | Anxiety: Generalized Anxiety Disorder | 15 | 0.1 |
| cTAKES-SDoH | entailment-RoBERTa | Anxiety: hx of anxiety state | 0 | 1 |
| cTAKES-SDoH | entailment-RoBERTa | Anxiety: Signs and symptoms of anxiety | 26 | 5e-04 |
| cTAKES-SDoH | entailment-RoBERTa | Depression: Family hx: Depression | -19 | 0.1 |
| cTAKES-SDoH | entailment-RoBERTa | Depression: hx of Depression | 4 | 0.7 |
| cTAKES-SDoH | entailment-RoBERTa | Depression: Major depressive disorder | 16 | 0.01 |
| cTAKES-SDoH | entailment-RoBERTa | Depression: Symptoms of depression | 39 | 2e-04 |
| cTAKES-SDoH | entailment-RoBERTa | Financial_strain: Financial problem | 49 | 0.04 |
| cTAKES-SDoH | entailment-RoBERTa | Financial_strain: NA | 35 | 0.1 |
| cTAKES-SDoH | entailment-RoBERTa | Housing: Homeless | 43 | 0.3 |
| cTAKES-SDoH | entailment-RoBERTa | Insurance_status: NA | 38 | 0.03 |
| cTAKES-SDoH | entailment-RoBERTa | Marital_or_partnership_status: Divorced | -22 | 0.01 |
| cTAKES-SDoH | entailment-RoBERTa | Marital_or_partnership_status: Married | 23 | 6e-05 |
| cTAKES-SDoH | entailment-RoBERTa | Marital_or_partnership_status: NA | 43 | 0.05 |
| cTAKES-SDoH | entailment-RoBERTa | Marital_or_partnership_status: Partner | -3 | 0.8 |
| cTAKES-SDoH | entailment-RoBERTa | Marital_or_partnership_status: Partner relationship problem | 17 | 0.3 |
| cTAKES-SDoH | entailment-RoBERTa | Marital_or_partnership_status: Single person | 13 | 0.05 |
| cTAKES-SDoH | entailment-RoBERTa | Marital_or_partnership_status: Widowed | 31 | 0.03 |
| cTAKES-SDoH | entailment-RoBERTa | Social_support: Has social support | 51 | 6e-09 |
| cTAKES-SDoH | entailment-RoBERTa | Social_support: Lives alone | 25 | 0.01 |
| cTAKES-SDoH | entailment-RoBERTa | Social_support: Lives with | 61 | 4e-08 |
| cTAKES-SDoH | entailment-RoBERTa | Social_support: NA | 21 | 0.2 |
| cTAKES-SDoH | entailment-RoBERTa | Social_support: Personal relationship breakdown | 6 | 0.4 |
| cTAKES-SDoH | entailment-RoBERTa | Transportation: Has access to a car | 53 | 4e-06 |
| cTAKES-SDoH | entailment-RoBERTa | Transportation: Has access to public transport vehicle | 57 | 6e-05 |
| cTAKES-SDoH | entailment-RoBERTa | Transportation: Transportation problems | 27 | 0.06 |
| CNN | hybrid | Anxiety: family hx: Anxiety state | 33 | 0.3 |
| CNN | hybrid | Anxiety: Generalized Anxiety Disorder | 17 | 0.2 |
| CNN | hybrid | Anxiety: hx of anxiety state | 2 | 0.7 |
| CNN | hybrid | Anxiety: NA | 4 | 0.5 |
| CNN | hybrid | Anxiety: Signs and symptoms of anxiety | 0 | 1 |
| CNN | hybrid | Depression: Family hx: Depression | 7 | 0.4 |
| CNN | hybrid | Depression: hx of Depression | 1 | 0.8 |
| CNN | hybrid | Depression: Major depressive disorder | 16 | 0.2 |
| CNN | hybrid | Depression: Symptoms of depression | 3 | 0.6 |
| CNN | hybrid | Housing: Homeless | -9 | 0.8 |
| CNN | hybrid | Housing: Stably housed | -6 | 0.3 |
| CNN | hybrid | Insurance_status: NA | 28 | 0.09 |
| CNN | hybrid | macro avg | 4 | 0.2 |
| CNN | hybrid | Marital_or_partnership_status: Divorced | -6 | 0.2 |
| CNN | hybrid | Marital_or_partnership_status: Married | -3 | 0.1 |
| CNN | hybrid | Marital_or_partnership_status: Partner | 2 | 0.8 |
| CNN | hybrid | Marital_or_partnership_status: Separated | 45 | 0.03 |
| CNN | hybrid | Marital_or_partnership_status: Single person | -5 | 0.07 |
| CNN | hybrid | Marital_or_partnership_status: Widowed | -53 | 3e-04 |
| CNN | hybrid | pain_and_disability: NA | 1 | 0.7 |
| CNN | hybrid | pain_and_disability: Pain intensity rating scale, current | 8 | 0.7 |
| CNN | hybrid | pain_and_disability: Pain intensity rating scale, worst | 18 | 0.3 |
| CNN | hybrid | Social_support: Has social support | 1 | 0.8 |
| CNN | hybrid | Social_support: Lives alone | -8 | 0.2 |
| CNN | hybrid | Social_support: Lives with | -17 | 0.007 |
| CNN | hybrid | Transportation: Has access to a car | 19 | 0.05 |
| CNN | hybrid | Transportation: Transportation problems | 2 | 0.9 |
| CNN | hybrid | weighted avg | 23 | 4e-05 |
| CNN | entailment-RoBERTa-mean | Anxiety: family hx: Anxiety state | 35 | 0.3 |
| CNN | entailment-RoBERTa-mean | Anxiety: Generalized Anxiety Disorder | -8 | 0.3 |
| CNN | entailment-RoBERTa-mean | Anxiety: hx of anxiety state | -28 | 0.01 |
| CNN | entailment-RoBERTa-mean | Anxiety: Signs and symptoms of anxiety | -20 | 8e-04 |
| CNN | entailment-RoBERTa-mean | Depression: Family hx: Depression | -34 | 0.02 |
| CNN | entailment-RoBERTa-mean | Depression: hx of Depression | -53 | 5e-04 |
| CNN | entailment-RoBERTa-mean | Depression: Major depressive disorder | -2 | 0.8 |
| CNN | entailment-RoBERTa-mean | Depression: Symptoms of depression | -26 | 8e-05 |
| CNN | entailment-RoBERTa-mean | Housing: Stably housed | -17 | 0.005 |
| CNN | entailment-RoBERTa-mean | Insurance_status: NA | 21 | 0.2 |
| CNN | entailment-RoBERTa-mean | Marital_or_partnership_status: Divorced | -33 | 0.001 |
| CNN | entailment-RoBERTa-mean | Marital_or_partnership_status: Married | 0 | 0.9 |
| CNN | entailment-RoBERTa-mean | Marital_or_partnership_status: Partner | -39 | 9e-04 |
| CNN | entailment-RoBERTa-mean | Marital_or_partnership_status: Single person | -10 | 0.003 |
| CNN | entailment-RoBERTa-mean | Marital_or_partnership_status: Widowed | -13 | 0.1 |
| CNN | entailment-RoBERTa-mean | Social_support: Has social support | -3 | 0.6 |
| CNN | entailment-RoBERTa-mean | Social_support: Lives alone | 13 | 0.06 |
| CNN | entailment-RoBERTa-mean | Social_support: Lives with | 10 | 0.08 |
| CNN | entailment-RoBERTa-mean | Transportation: Has access to a car | 53 | 8e-07 |
| CNN | entailment-RoBERTa-mean | Transportation: Transportation problems | -23 | 0.2 |
| CNN | entailment-RoBERTa | Anxiety: family hx: Anxiety state | 2 | 0.9 |
| CNN | entailment-RoBERTa | Anxiety: Generalized Anxiety Disorder | 0 | 1 |
| CNN | entailment-RoBERTa | Anxiety: hx of anxiety state | -27 | 0.01 |
| CNN | entailment-RoBERTa | Anxiety: Signs and symptoms of anxiety | -32 | 1e-04 |
| CNN | entailment-RoBERTa | Depression: Family hx: Depression | -36 | 0.02 |
| CNN | entailment-RoBERTa | Depression: hx of Depression | -38 | 0.02 |
| CNN | entailment-RoBERTa | Depression: Major depressive disorder | 11 | 0.1 |
| CNN | entailment-RoBERTa | Depression: Symptoms of depression | -12 | 0.08 |
| CNN | entailment-RoBERTa | Housing: Stably housed | -1 | 0.7 |
| CNN | entailment-RoBERTa | Insurance_status: NA | 21 | 0.2 |
| CNN | entailment-RoBERTa | Marital_or_partnership_status: Divorced | -32 | 0.001 |
| CNN | entailment-RoBERTa | Marital_or_partnership_status: Married | 0 | 1 |
| CNN | entailment-RoBERTa | Marital_or_partnership_status: Partner | -43 | 5e-04 |
| CNN | entailment-RoBERTa | Marital_or_partnership_status: Single person | -16 | 2e-05 |
| CNN | entailment-RoBERTa | Marital_or_partnership_status: Widowed | -16 | 0.06 |
| CNN | entailment-RoBERTa | Social_support: Has social support | 22 | 0.009 |
| CNN | entailment-RoBERTa | Social_support: Lives alone | 7 | 0.3 |
| CNN | entailment-RoBERTa | Social_support: Lives with | 2 | 0.7 |
| CNN | entailment-RoBERTa | Transportation: Has access to a car | 43 | 5e-06 |
| CNN | entailment-RoBERTa | Transportation: Transportation problems | -40 | 0.06 |
| hybrid | entailment-RoBERTa-mean | Anxiety: family hx: Anxiety state | 15 | 0.6 |
| hybrid | entailment-RoBERTa-mean | Anxiety: Generalized Anxiety Disorder | -25 | 0.05 |
| hybrid | entailment-RoBERTa-mean | Anxiety: hx of anxiety state | -30 | 0.009 |
| hybrid | entailment-RoBERTa-mean | Anxiety: Signs and symptoms of anxiety | -21 | 0.006 |
| hybrid | entailment-RoBERTa-mean | Depression: Family hx: Depression | -41 | 0.01 |
| hybrid | entailment-RoBERTa-mean | Depression: hx of Depression | -54 | 5e-04 |
| hybrid | entailment-RoBERTa-mean | Depression: Major depressive disorder | -17 | 0.1 |
| hybrid | entailment-RoBERTa-mean | Depression: Symptoms of depression | -29 | 1e-04 |
| hybrid | entailment-RoBERTa-mean | Financial_strain: Financial problem | 24 | 0.3 |
| hybrid | entailment-RoBERTa-mean | Financial_strain: NA | 8 | 0.7 |
| hybrid | entailment-RoBERTa-mean | Housing: Stably housed | -11 | 0.03 |
| hybrid | entailment-RoBERTa-mean | Housing: Subsidized housing | 35 | 0.07 |
| hybrid | entailment-RoBERTa-mean | Insurance_status: NA | -7 | 0.7 |
| hybrid | entailment-RoBERTa-mean | Marital_or_partnership_status: Divorced | -27 | 0.004 |
| hybrid | entailment-RoBERTa-mean | Marital_or_partnership_status: Married | 3 | 0.08 |
| hybrid | entailment-RoBERTa-mean | Marital_or_partnership_status: NA | 22 | 0.3 |
| hybrid | entailment-RoBERTa-mean | Marital_or_partnership_status: Partner | -42 | 0.001 |
| hybrid | entailment-RoBERTa-mean | Marital_or_partnership_status: Partner relationship problem | -5 | 0.8 |
| hybrid | entailment-RoBERTa-mean | Marital_or_partnership_status: Separated | -54 | 0.02 |
| hybrid | entailment-RoBERTa-mean | Marital_or_partnership_status: Single person | -5 | 0.1 |
| hybrid | entailment-RoBERTa-mean | Marital_or_partnership_status: Widowed | 40 | 0.002 |
| hybrid | entailment-RoBERTa-mean | Social_support: Has social support | -4 | 0.1 |
| hybrid | entailment-RoBERTa-mean | Social_support: Lives alone | 21 | 0.002 |
| hybrid | entailment-RoBERTa-mean | Social_support: Lives with | 27 | 4e-05 |
| hybrid | entailment-RoBERTa-mean | Transportation: Has access to a car | 34 | 0.004 |
| hybrid | entailment-RoBERTa-mean | Transportation: Has access to public transport vehicle | 31 | 0.01 |
| hybrid | entailment-RoBERTa-mean | Transportation: Transportation problems | -25 | 0.2 |
| hybrid | entailment-RoBERTa | Anxiety: family hx: Anxiety state | -18 | 0.5 |
| hybrid | entailment-RoBERTa | Anxiety: Generalized Anxiety Disorder | -17 | 0.2 |
| hybrid | entailment-RoBERTa | Anxiety: hx of anxiety state | -29 | 0.01 |
| hybrid | entailment-RoBERTa | Anxiety: Signs and symptoms of anxiety | -32 | 6e-04 |
| hybrid | entailment-RoBERTa | Depression: Family hx: Depression | -43 | 0.007 |
| hybrid | entailment-RoBERTa | Depression: hx of Depression | -40 | 0.02 |
| hybrid | entailment-RoBERTa | Depression: Major depressive disorder | -4 | 0.7 |
| hybrid | entailment-RoBERTa | Depression: Symptoms of depression | -15 | 0.04 |
| hybrid | entailment-RoBERTa | Financial_strain: Financial problem | 40 | 0.1 |
| hybrid | entailment-RoBERTa | Financial_strain: NA | 22 | 0.4 |
| hybrid | entailment-RoBERTa | Housing: Stably housed | 4 | 0.3 |
| hybrid | entailment-RoBERTa | Housing: Subsidized housing | 56 | 0.02 |
| hybrid | entailment-RoBERTa | Insurance_status: NA | -7 | 0.7 |
| hybrid | entailment-RoBERTa | Marital_or_partnership_status: Divorced | -26 | 0.004 |
| hybrid | entailment-RoBERTa | Marital_or_partnership_status: Married | 3 | 0.09 |
| hybrid | entailment-RoBERTa | Marital_or_partnership_status: NA | 22 | 0.3 |
| hybrid | entailment-RoBERTa | Marital_or_partnership_status: Partner | -45 | 7e-04 |
| hybrid | entailment-RoBERTa | Marital_or_partnership_status: Partner relationship problem | -1 | 0.9 |
| hybrid | entailment-RoBERTa | Marital_or_partnership_status: Separated | -49 | 0.03 |
| hybrid | entailment-RoBERTa | Marital_or_partnership_status: Single person | -11 | 0.003 |
| hybrid | entailment-RoBERTa | Marital_or_partnership_status: Widowed | 38 | 0.003 |
| hybrid | entailment-RoBERTa | Social_support: Has social support | 20 | 5e-05 |
| hybrid | entailment-RoBERTa | Social_support: Lives alone | 15 | 0.01 |
| hybrid | entailment-RoBERTa | Social_support: Lives with | 19 | 8e-04 |
| hybrid | entailment-RoBERTa | Transportation: Has access to a car | 24 | 0.02 |
| hybrid | entailment-RoBERTa | Transportation: Has access to public transport vehicle | 28 | 0.02 |
| hybrid | entailment-RoBERTa | Transportation: Transportation problems | -43 | 0.05 |
| entailment-RoBERTa-mean | entailment-RoBERTa | Anxiety: family hx: Anxiety state | -33 | 0.2 |
| entailment-RoBERTa-mean | entailment-RoBERTa | Anxiety: Generalized Anxiety Disorder | 8 | 0.4 |
| entailment-RoBERTa-mean | entailment-RoBERTa | Anxiety: hx of anxiety state | 1 | 0.9 |
| entailment-RoBERTa-mean | entailment-RoBERTa | Anxiety: Level of anxiety | -1 | 0.9 |
| entailment-RoBERTa-mean | entailment-RoBERTa | Anxiety: Signs and symptoms of anxiety | -11 | 0.04 |
| entailment-RoBERTa-mean | entailment-RoBERTa | Depression: Family hx: Depression | -2 | 0.9 |
| entailment-RoBERTa-mean | entailment-RoBERTa | Depression: hx of Depression | 14 | 0.3 |
| entailment-RoBERTa-mean | entailment-RoBERTa | Depression: Major depressive disorder | 13 | 0.03 |
| entailment-RoBERTa-mean | entailment-RoBERTa | Depression: Symptoms of depression | 14 | 0.04 |
| entailment-RoBERTa-mean | entailment-RoBERTa | Financial_strain: Financial problem | 15 | 0.5 |
| entailment-RoBERTa-mean | entailment-RoBERTa | Financial_strain: Financially secure | 8 | 0.7 |
| entailment-RoBERTa-mean | entailment-RoBERTa | Financial_strain: NA | 14 | 0.5 |
| entailment-RoBERTa-mean | entailment-RoBERTa | Food: NA | 0 | 1 |
| entailment-RoBERTa-mean | entailment-RoBERTa | Food: Provision of food | 20 | 0.7 |
| entailment-RoBERTa-mean | entailment-RoBERTa | Housing: Homeless | 42 | 0.1 |
| entailment-RoBERTa-mean | entailment-RoBERTa | Housing: Marginally housed | -2 | 0.7 |
| entailment-RoBERTa-mean | entailment-RoBERTa | Housing: Stably housed | 15 | 2e-05 |
| entailment-RoBERTa-mean | entailment-RoBERTa | Housing: Subsidized housing | 21 | 0.08 |
| entailment-RoBERTa-mean | entailment-RoBERTa | Insurance_status: NA | 0 | 1 |
| entailment-RoBERTa-mean | entailment-RoBERTa | Marital_or_partnership_status: Divorced | 1 | 0.9 |
| entailment-RoBERTa-mean | entailment-RoBERTa | Marital_or_partnership_status: Engaged to be married | 0 | 1 |
| entailment-RoBERTa-mean | entailment-RoBERTa | Marital_or_partnership_status: Married | 0 | 0.9 |
| entailment-RoBERTa-mean | entailment-RoBERTa | Marital_or_partnership_status: NA | 0 | 1 |
| entailment-RoBERTa-mean | entailment-RoBERTa | Marital_or_partnership_status: Partner | -4 | 0.5 |
| entailment-RoBERTa-mean | entailment-RoBERTa | Marital_or_partnership_status: Partner relationship problem | 3 | 0.9 |
| entailment-RoBERTa-mean | entailment-RoBERTa | Marital_or_partnership_status: Separated | 4 | 0.4 |
| entailment-RoBERTa-mean | entailment-RoBERTa | Marital_or_partnership_status: Single person | -6 | 0.04 |
| entailment-RoBERTa-mean | entailment-RoBERTa | Marital_or_partnership_status: Widowed | -3 | 0.7 |
| entailment-RoBERTa-mean | entailment-RoBERTa | Social_support: Has social support | 25 | 3e-06 |
| entailment-RoBERTa-mean | entailment-RoBERTa | Social_support: Lives alone | -6 | 0.2 |
| entailment-RoBERTa-mean | entailment-RoBERTa | Social_support: Lives with | -8 | 0.08 |
| entailment-RoBERTa-mean | entailment-RoBERTa | Social_support: NA | 0 | 1 |
| entailment-RoBERTa-mean | entailment-RoBERTa | Social_support: Personal relationship breakdown | 2 | 0.8 |
| entailment-RoBERTa-mean | entailment-RoBERTa | Social_support: Social Support | -3 | 0.9 |
| entailment-RoBERTa-mean | entailment-RoBERTa | Transportation: Has access to a car | -9 | 0.06 |
| entailment-RoBERTa-mean | entailment-RoBERTa | Transportation: Has access to public transport vehicle | -4 | 0.7 |
| entailment-RoBERTa-mean | entailment-RoBERTa | Transportation: NA | -3 | 0.9 |
| entailment-RoBERTa-mean | entailment-RoBERTa | Transportation: Transportation problems | -17 | 0.1 |

Supplementary Table 9. ANOVA of factors contributing to F_1_ model performance

| **term** | **degrees of freedom** | **sumsq** | **meansq** | **statistic** | **p-value** |
| --- | --- | --- | --- | --- | --- |
| label | 9 | 16 | 1.78 | 83.6 | 1.08e-65 |
| fold | 1 | 0.00172 | 0.00172 | 0.0808 | 0.776 |
| method | 3 | 4.41 | 1.47 | 68.9 | 2.22e-31 |
| Residuals | 218 | 4.65 | 0.0213 | NA | NA |

Supplementary Table 10. Correlation of per-label F_1_ between methods

| **model 1** | **model 2** | **Pearson R** | **p-value** |
| --- | --- | --- | --- |
| cTAKES-SDoH | hybrid | 0.536 | 1.2e-05 |
| cTAKES-SDoH | CNN | 0.607 | 3.4e-07 |
| cTAKES-SDoH | entailment-RoBERTA | 0.787 | 2.92e-12 |
| hybrid | CNN | 0.929 | 2.59e-26 |
| hybrid | entailment-RoBERTA | 0.682 | 1.33e-08 |
| CNN | entailment-RoBERTA | 0.819 | 6.49e-14 |

Supplementary Table 11. Qualitative analysis of entailment model sensitivity to wording

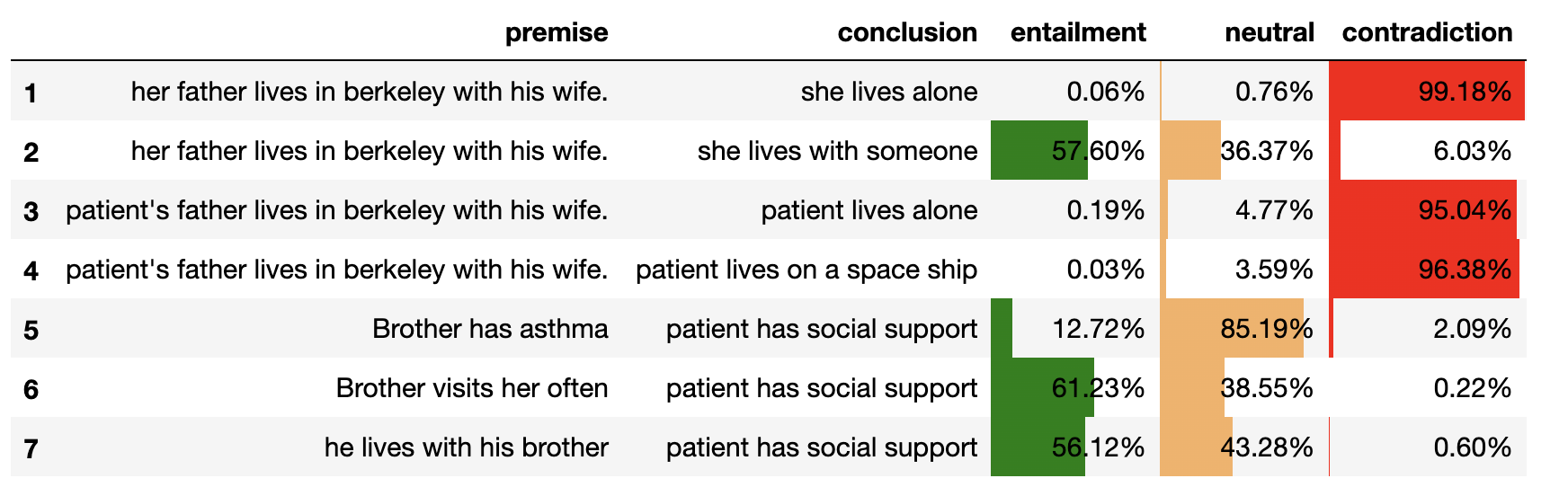

### **Supplementary Methods**

Analysis of Manual Annotations

*Computation of weighted Cohen’s kappa*

For each pair of annotators (*a* and *b*) and each label of class *k*, kappa κ*_k_^ab^* was calculated together with an average number of labels of class *k* present in annotations by *a* and *b* (*n_k_^ab^*), and the weighted average per label was obtained as:

$$\kappa_{k}=\frac{\sum_{ab} \kappa_{k}^{ab}\cdot n_{k}^{ab}}{\sum_{ab} n_{k}^{ab}}$$

*Choice of inter-rater agreement method for NER*

For scoring inter-rater agreement, Cohen’s kappa is a common metric []. However, in the domain of NER, its application is challenging because (1) Cohen’s kappa takes into consideration true negatives and (2) as in NER task true negatives are normally abundant and may dominate the estimate, and (3) negatives are not tracked in common data structures for NER. To understand condition (2), consider that a note may consist of 2—4 thousands of tokens, with only about 5—10 tokens belonging to named entities. Thus, all tokens that don't belong to any entity and predicted as such will contribute with the same weight as true positive predictions. On the other hand, F_1_ does not account for true negatives.

cTAKES configuration

cTAKES, designed for the task of extracting structured information from clinical text, has its roots in 2006 at the Mayo Clinic where a team was led by Dr. Guergana Savova and Dr. [Christopher Chute](https://en.wikipedia.org/wiki/Christopher_G._Chute). Since 2012 it has been a top-level Apache project. The project was designed around the task of extracting structured information from clinical text.

cTAKES functionality involves not only identifying and coding clinical concepts and other named entities, but also extracting relationships and concept modifiers such as negation detection, degree-of-detection, and co-referencing.   Over the years the project has been expanded and improved by contributors from various centers of excellence in the domains of medicine and informatics including Boston Children’s Hospital, the Cleveland Clinic, and Harvard School of Medicine.

Rather than creating a monolithic application with the risk of an ever-expanding complexity and instability, the authors designed the product around the UIMA text processing framework in which configuration builds a pipeline of stages through which the processing proceeds.   Interim results are stored in an object called the CAS, an acronym for a rather generically titled “Common Analysis System”.   Each step can create Annotations and have visibility into any Annotations that were created by earlier stages in the process.  Not commonly used is a feature of the CAS that could house multiple notes, aka Sofa objects, each with its own extracted data and an ability to query information from sibling notes.  UIMA Annotations are configurable data structures defined in a Type System that leads to the automatic generation of Java code which implements the Types used.

A typical Annotation is a data structure that references a Sofa text object, start and end offsets relative to that text, and attributes specific to the annotation.   Some Annotators only add attributes to a pre-existing Annotation. For instance, the Negex annotator sets the value of the attribute “polarity” in a subtype of IdentifiedAnnotation, such as MedicationMention or SignSymptomMention.   Polarity reflects whether the concept has been identified as negated in the text:  as in “no fever”, or “denies headache”.

Most of the annotators have support files that range from simple lists to weight vectors created by ML pre-processing.

A pipeline usually begins with annotators that deconstruct the text into sentences, chunks, and semantic tokens.  Once these generic annotations are in place, they can be used by domain focused annotators.

One of the core annotators is dedicated to concept identification using the UMLS distribution of one or more vocabularies.  These vocabularies usually include SNOMED, RXNORM, and a meta concept vocabulary where terms are identified by CUIs (concept unique identifiers).   ICD10 and other pertinent vocabularies can be brought into the dictionary package.

Over the last 15 years, cTAKES has been applied to many tasks, using custom pipelines and Annotators built to accomplish the goals of each project in the most efficient manner.  Users have creatively inserted cTAKES into their overall information strategy in areas that range from retrospective demographic research to point-of-care decision support.

The core framework has also been improved so that the pipeline can be determined by a single configuration file that invokes the initialization code resident in each of the Annotation modules.   But generally, any production use of cTAKES will require significant programming and information transformation skills.

From a technical perspective, the UCSF Information Commons implementation of cTAKES repackaged the application as a web service application server.  This allows it to be completely decoupled from the design of either upstream or downstream data persistence stages.

Note texts are passed to these web services one at a time and their return message depends upon the service that was invoked.  One API returns a serialized dump of the entire CAS object.  Another, designed for quick incorporation into research, synthesizes and groups a set of concept objects in clinical domains, pulling information from multiple Annotation types in the CAS to synthesize each Concept object.

The entire distribution consists of over two thousand Java source files comprised of several hundreds of thousands of lines of code. To ease uptake of the application, the cTAKES team has created a running base configuration consisting of a minimal pipeline and UMLS vocabulary.

We use this default configuration as a baseline in evaluating the success of cTAKES at fulfilling the objectives of the SDoH configuration efforts. For this configuration we will be using the default dictionary known as SnoRX_ab.  Our default pipeline includes these annotators:

   add SimpleSegmentAnnotator

   org.apache.cTAKES.core.ae.SentenceDetector

   add org.apache.cTAKES.core.ae.TokenizerAnnotatorPTB

   add org.apache.cTAKES.contexttokenizer.ae.ContextDependentTokenizerAnnotator

   addDescription POSTagger

   add Chunker

   addDescription adjuster.ChunkAdjuster NP,NP 1

   addDescription adjuster.ChunkAdjuster NP,PP,NP 2

   add DefaultJCasTermAnnotator LookupXml=org/apache/cTAKES/dictionary/lookup/fast/sno_rx_16ab.xml

   addDescription ClearNLPDependencyParserAE

   add org.apache.cTAKES.dependency.parser.ae.ClearNLPSemanticRoleLabelerAE

   package org.apache.cTAKES.assertion.medfacts.cleartk

   addDescription PolarityCleartkAnalysisEngine

   addDescription HistoryCleartkAnalysisEngine

   addDescription SubjectCleartkAnalysisEngine

   add LocationOfRelationExtractorAnnotator

As a step towards the current implementation of the SDoH configuration, we moved to the configuration created for the InfoCommons bulk concept extraction project.  Here, some 100 million notes across multiple medical venues were parsed and made available to researchers at UCSF. This pipeline included an improved negation annotator, code to look for “location of” attributes, and the addition of case sensitive and case insensitive black-list files that prevent annotation of concepts that are unneeded, or rarely used acronyms.   The dictionary is newly extracted from the UMLS 2020 release and includes clinical, nursing, and some behavioral terms as well as a gene vocabulary.  Concepts are mapped to their HGNC, RXNORM, and SNOMED codes   The new dictionary is quite large, but it needed to be further customized so a small package of scripts was created to manage the differences between the new baseline and the customized baseline.   We discovered that there were several overlaps between certain acronyms for clinical concepts and gene symbols. These caused many spurious identified concepts to be generated.

The InfoCommons Pipeline consists of these annotators

   add SimpleSegmentAnnotator

   add  SentenceDetectorAnnotatorBIO classifierJarPath=/org/apache/cTAKES/core/sentdetect/model.jar

   add org.apache.cTAKES.core.ae.TokenizerAnnotatorPTB

   add org.apache.cTAKES.contexttokenizer.ae.ContextDependentTokenizerAnnotator

   addDescription POSTagger

   add Chunker

   addDescription adjuster.ChunkAdjuster NP,NP 1

   addDescription adjuster.ChunkAdjuster NP,PP,NP 2

   add org.apache.cTAKES.dictionary.lookup2.ae.OverlapJCasTermAnnotator LookupXml=org/apache/cTAKES/dictionary/lookup/fast/ucsf_dict_v3.xml minimumSpan=2

   addDescription ClearNLPDependencyParserAE

   add LabValueFinder labTUIs=T028,T192,T116

   add org.apache.cTAKES.ytex.uima.annotators.NegexAnnotator

   add org.apache.cTAKES.dependency.parser.ae.ClearNLPSemanticRoleLabelerAE

   package org.apache.cTAKES.assertion.medfacts.cleartk

   addDescription UncertaintyCleartkAnalysisEngine

   addDescription HistoryCleartkAnalysisEngine

   addDescription ConditionalCleartkAnalysisEngine

   addDescription SubjectCleartkAnalysisEngine

   add LocationOfRelationExtractorAnnotator classifierJarPath=/org/apache/cTAKES/relationextractor/models/location_of/model.jar

Finally, we implemented a further customization of the InfoCommons pipeline for the SDoH purposes. Here, the dictionary of InfoCommons was augmented with social risk factor and QOL terms.  We removed the genetic information from the dictionary since it would not have been useful in this study and just bloated the dictionary.  In addition, the following was added or changed:

- Based on example notes we had seen, we added more synonyms of existing concepts to the areas of back pain, depression, anxiety, and social and financial risk factors.
- As we needed to record pain and anxiety scores where they occurred in the notes, we added to the pipeline a new annotator, TokensRegex.  This allowed us to create regular expressions both on text as well as on semantic or linguistic token types or on the lemmas of a given word.  These could capture phrases such as “down to 5” or “worse in the morning at 7-8”  where the term pain might not even be present.
- We added a custom-built PredicateAnnotator to the pipeline that looked for phrases acting as predicates to coded concepts.  In some cases, this would help locate text that gave qualitative or quantitative context to an IdentifiedAnnotation.  For example, in the sentence, “reports back pain *intensifying particularly at night*”, the Annotator would capture the italicized words in association with the code-able concept of back pain, the idea being that the predicate could then be looked at by an ML or other textual analysis step, post cTAKES.

The SDoH pipeline consists of the following stages.

   add SimpleSegmentAnnotator

   add org.apache.cTAKES.core.ae.SentenceDetector

   add org.apache.cTAKES.core.ae.TokenizerAnnotatorPTB

   add org.apache.cTAKES.contexttokenizer.ae.ContextDependentTokenizerAnnotator

   addDescription POSTagger

   add Chunker

   addDescription adjuster.ChunkAdjuster NP,NP 1

   addDescription adjuster.ChunkAdjuster NP,PP,NP 2

   add org.apache.cTAKES.dictionary.lookup2.ae.OverlapJCasTermAnnotator LookupXml=org/apache/cTAKES/dictionary/lookup/fast/cocoa_v1.xml minimumSpan=2

   addDescription ClearNLPDependencyParserAE

   add org.apache.cTAKES.dependency.parser.ae.ClearNLPSemanticRoleLabelerAE

   package org.apache.cTAKES.assertion.medfacts.cleartk

   addDescription UncertaintyCleartkAnalysisEngine

   addDescription HistoryCleartkAnalysisEngine

   addDescription ConditionalCleartkAnalysisEngine

   addDescription SubjectCleartkAnalysisEngine

   add org.apache.cTAKES.ytex.uima.annotators.NegexAnnotator

   add org.apache.cTAKES.constituency.parser.ae.ConstituencyParser

   add ModifierExtractorAnnotator classifierJarPath=/org/apache/cTAKES/relationextractor/models/modifier_extractor/model.jar

   add LocationOfRelationExtractorAnnotator classifierJarPath=/org/apache/cTAKES/relationextractor/models/location_of/model.jar

   addDescription org.ucsf.cTAKES.tkregex.ae.TokensRegexNER TokenRegexRules=resources/tkregex/cocoatk2.txt

   add org.ucsf.cTAKES.predicatefinder.ae.PredicateFinder PhraseTops="VB,NVP,TOP,INC,FRAG,S,S-RED,NP" PredicateLeaves="NP-ADV,NP,VP,NP-PRD,PP,NP-TMP,ADVP-TMP" MentionTypes="SignSymptomMention,ProcedureMention,DiseaseDisorderMention,MedicationMention"

### **Supplementary Results**

**Qualitative analysis of cTAKES predictions**

Pain and disability scores were often missed while cTAKES captured score names (“VAS”) but not values. On the other hand, most false positives (resulting in a first-level precision of 42.87%) arose due to factors such as misinterpretation of terms from other medical domains, e.g., “separation” (from an intimate partner) interpreted as “diastasis,” “bf” for “boyfriend” interpreted as “breast feeding”, “cab” interpreted as “coronary artery bypass surgery”, “IHSS” (an acronym for “in-home support services”) interpreted as “idiopathic hypertrophic subaortic stenosis”). Moreover, modifiers were occasionally missed (e.g., “ex” missed in “ex husband”), or modifiers from other lines of text were mistakenly assigned to the entity of interest. Some SDoH were falsely captured in empty questionnaire fields, e.g., “Relationship status:” followed by “Not on file” or by a blank.

**Qualitative evaluation of RoBERTA entailment model**

We saw that both the BoW and the CNN models had difficulty detecting entities due to fundamental limitations of BoW method, and potentially low sample size in case of the CNN model. Therefore, we attempted to qualitatively assess the potential of more sophisticated models designed and earlier trained (without fine-tuning on our dataset) on a selected passages that appeared most challenging for BoW and CNN models. The results appear promising (Fig. 4, main text, Supplemental Fig 10) in that the model can detect alternative phrasings of relevant events in most cases, with few false positives cases.

***Detection of implication***

For example, the model correctly concludes that “She lives in section 8 housing” entails “Patient has low income”.

“Not being able to afford medical cannabis juice” is detected as entailing that “Patient struggles financially”

“grief (due to recent passing of husband)” is detected as “patient is widowed”, but mentions of fiancé / fiancée are not detected as “patient is engaged to be married”.

***Errors due to wrong subject reference resolution.***

For example, referring to the subject as “patient” instead of by a 3^rd^ person singular pronoun (she/he) may degrade prediction score by 2—10%. Additionally, the entailment model fails on several examples where multiple subjects are mentioned (e.g., “*Patient's father lives ... with his wife*” is deemed to entail that “*Patient lives with someone*” and to contradict with “*Patient lives alone*”).

Multiple cases of mentions of a patient living with a spouse and a child are deemed imply that “This describes the marital status of his / her parents”.

***Sensitivity to predicate phrasing***

Precise wording may matter, as for example a premise “*grief (due to recent passing of husband)*” is deemed to entail that “*she feels down*” but not that “*she is depressed*”. On the other hand, a sentence “*She presents … with … thoughts of hopelessness*” triggers entailment with both indicated hypotheses, albeit with a higher probability for “*down*” compared to “*depressed*”.

References of patient driving a truck or a van are deemed to imply that “Patient drives a vehicle”, but not “Patient drives a car”.

***False positives***

“He is desperate to get back to work” is deemed to contradict that “The patient is employed”, even though it may mean either lack of employment or desire to return to work from a sick leave.

Multiple cases of prescription instructions (“… take by mouth…”) are deemed to imply “This provides information about patient’s nutrition”.
